# Trajectories of plasma biomarkers, amyloid-beta burden and cognitive decline in Alzheimer’s disease: A Longitudinal ADNI Study

**DOI:** 10.1101/2025.09.30.25337003

**Authors:** Yara Yakoub, Ting Qiu, Clémence Peyrot, Gemma Salvadó, Sylvia Villeneuve, Alexa Pichet Binette, Alzheimer’s Disease Neuroimaging Initiative (ADNI)

**Affiliations:** Douglas Mental Health University Institute, Montreal, Quebec, Canada Address: 6875 Blvd. LaSalle, Verdun, H4H 1R3, Québec, Canada; Integrated Program in Neuroscience, Faculty of medicine, McGill University, Montreal, Canada Address: 1033 Pine Ave. W. Montreal, H3A 1A1, Québec, Canada; Centre de Recherche de l’Institut Universitaire de Gériatrie de Montréal, Montréal, Québec, Canada Address: 4565 Queen Mary Rd, Montreal, H3W 1W5, Quebec, Canada; Barcelonaβeta Brain Research Centre (BBRC), Pasqual Maragall Foundation, Barcelona, Spain Address: Carrer de Wellington, 30, Sant Martí, 08005 Barcelona, Spain; Hospital del Mar Research Institute, Barcelona, Spain Address: Carrer del Doctor Aiguader, 88, Ciutat Vella, 08003 Barcelona, Spain; Clinical Memory Research Unit, Department of Clinical Sciences Malmö, Lund University, Lund, Sweden Address: S:t Johannesgatan 8 211 46 Malmö Sweden; Montreal Neurological Institute, McGill University, Montreal, Canada Address: 3801 Rue University, Ville-Marie, Montreal, H3A 2B4, Québec, Canada; Department of Psychiatry, Faculty of medicine, McGill University, Montreal, Canada Address: 1033 Pine Ave. W. Montreal, H3A 1A1, Québec, Canada; Department of Physiology and Pharmacology, Université de Montréal, Montréal, Quebec, Canada Address: Pavillon Roger-Gaudry, 2900 Edouard Montpetit Blvd, Montreal, H3T 1J4 Québec, Canada

## Abstract

As novel amyloid-β targeted therapies emerge, plasma biomarkers have promising potential to serve as screening tools and as surrogate measures for treatment outcomes. Understanding longitudinal trajectories of these biomarkers and how their changes relate to changes in AD pathology and cognition is needed to help track treatment response and guide patient care. We analyzed data from 394 individuals in the ADNI-FNIH dataset who had plasma biomarkers available across 14 assays, Aβ-PET scans and cognitive assessments over a 10-year period. Plasma p-tau217, regardless of the assay used, had the greatest rate of change over time. This increase was related to concurrent increase in Aβ-PET burden only in individuals with low levels of Aβ. The rate of p-tau217 change, rather than its baseline level, was the strongest predictor of future Aβ-PET positivity. On the other hand, in individuals with elevated levels of Aβ, higher rate of change in p-tau217 was associated with faster cognitive decline. These findings highlight a “dual” role of plasma p-tau217 rate of change, being either predictive of accumulating Aβ pathology at early stages and of cognitive decline at later stages of the AD continuum.

## INTRODUCTION

Plasma biomarkers have emerged as powerful tools for detecting Alzheimer’s disease (AD) pathology. As they become increasingly available worldwide, growing initiatives are establishing guidelines of practical use and acceptable performance, paving the way for clinical practice.^1-3^ Among the currently available biomarkers, plasma phosphorylated (p)-tau217 has shown strong potential as a reliable and clinically valuable marker, with high diagnostic accuracy when validated against established reference standards to measure amyloid-β (Aβ) pathology (autopsy/cerebrospinal fluid (CSF) /positron emission tomography (PET)).^4-10^ Cross-sectional plasma p-tau217 levels are also related to clinical progression and cognitive decline.^11, 12^ Other variants of p-tau markers (e.g. p-tau181, p-tau231) and plasma Aβ peptides (Aβ42/40) have also shown associations with AD related pathologies.^13-16^ Their diagnostic performance, however, was lower compared to p-tau217.^17, 18^ Other plasma markers like glial fibrillary acidic protein (GFAP) and neurofilament light (NfL) exhibit lower specificity for AD related pathologies.^19-22^

To date, most plasma biomarker studies in AD have been cross-sectional, with limited data on how these markers evolve longitudinally and how well they track clinically relevant outcomes.^13, 23-25^ In particular, evidence remains scarce on whether within-person changes in plasma biomarkers reliably mirror changes in insoluble Aβ pathology measured in vivo with PET, or whether they capture cognitive decline along the AD continuum.^26, 27^ Addressing this gap is critical as if plasma biomarker trajectories can serve as robust surrogates of disease progression, they could transform both the monitoring of anti-amyloid therapies and risk of progression. Establishing the longitudinal precision of these assays, and directly comparing how their rates of change relate to gold-standard outcomes (PET) and cognitive decline, is therefore a crucial step toward their clinical implementation, both for early prediction of cortical Aβ accumulation and at later disease stages for monitoring disease progression.^28-30^

In this study, we conducted the first large-scale head-to-head evaluation of longitudinal plasma biomarker trajectories within the Alzheimer’s Disease Neuroimaging Initiative, leveraging up to 10 years of follow-up in 394 participants from the FNIH biomarker consortium. We compared fourteen assays spanning Aβ42/40, p-tau217, p-tau181, NfL, and GFAP. We examined how their longitudinal changes related to concurrent Aβ accumulation and cognitive decline, as well as future Aβ-PET positivity. This comprehensive approach extends prior cross-sectional work by directly testing the pathophysiological and clinical relevance of change in plasma biomarkers across multiple platforms, offering unique insights into their utility as surrogate markers.

## RESULTS

### Study participants

On average, the 394 participants were 72.6 years old at the first plasma visit, 145 (36.8%) were Aβ-PET positive (defined as a Centiloid threshold above 20), 197 (50%) were women and 207 (52.5%) were cognitively unimpaired (**Table 1**). The average follow-up time for both plasma measurements and Aβ-PET scans was 5.5 years (SD = 1.7), with a range of 1.9 to 10.9 years. Participants had an average of three timepoints for both plasma and Aβ-PET. All analyses were performed across Aβ-PET groups and in the full sample. The Aβ-PET positive group was on average 2.67 years older and had 40% more participants with cognitive impairment than the Aβ-PET negative group (**Table 1**). Each biomarker was measured using multiple assays. P-tau217 and Aβ42/40 were measured with five assays and four assays, respectively, including the highest performing mass-spectrometry C2N assays (C2N p-tau217, %C2N p-tau217).^4, 31^ In this study, C2N P-tau217 and %p-tau were considered as separate measures within the comparison.

**Table 1.** Sample demographics. Mean (standard deviation), or where appropriate, count and percentage for both the full and subsample in the ADNI cohort. *P*-values were derived from Mann-Whitney U test for continuous variables and chi-squared for categorical variables. *Abbreviations*: F, female; APOE,apolipoprotein-E genotype; MMSE, Mini-Mental State Examination; MCI,Mild cognitive impairment.

| Characteristics | Full sample<br>(N = 394) | Aβ-Negative<br>(N = 249) | Aβ-Positive<br>(N = 145) | *P* value |
| --- | --- | --- | --- | --- |
| Age, years | 72.65 (7.12) | 71.67 (7.12) | 74.34 (6.81) | < 0.001 |
| Education (years) | 16.28 (2.69) | 16.56 (2.70) | 15.81 (2.62) | 0.004 |
| Sex (n, %) | 197 (50) | 123 (49.40) | 74 (51.03) | 0.83 |
| *APOE* ε4 carriers<br>(n, %) | 135 (34.26) | 57 (22.89) | 78 (53.79) | < 0.001 |
| Aβ-PET Centiloids | 23.71 (39.55) | -1.26 (10.80) | 66.59 (33.76) | < 0.001 |
| Aβ-PET change<br>(Centiloids/year) | 2.41 (1.94) | 1.46 (1.34) | 4.04 (1.74) | < 0.001 |
| MMSE (/30) | 28.58 (1.76) | 29.00 (1.29) | 27.85 (2.18) | < 0.001 |
| Clinical diagnosis<br>(n, %) |  |  |  | < 0.001 |
| Cognitively normal | 207 (52.54) | 146 (58.63) | 61 (42.07) |  |
| MCI | 180 (45.69) | 103 (41.37) | 77 (53.10) |  |
| AD dementia | 7 (1.78) | 0 (0) | 7 (4.83) |  |
| Follow-up time<br>(years) | 5.50 (1.74) | 5.76 (1.82) | 5.05 (1.51) |  |

These two biomarkers were the main focus of the study. Plasma p-tau181, NfL, and GFAP, each measured both with Roche Neurotoolkit and Quanterix’s Neurology 4-Plex, are presented in supplementary material. The correlations among plasma biomarker levels at baseline and their rate of change across the different platforms are found in **Fig. S1**.

### Longitudinal plasma biomarkers trajectories over time

First, we assessed how each plasma biomarker changed over time in the full sample as well as in the Aβ-PET positive and negative groups separately using linear mixed effect models. For In the whole group, plasma p-tau217 assays showed the significant steeper increase over time among all assays (β = 0.06 to 0.08, *p* < 0.001, **Fig. 1A**). There was a significant interaction between Aβ-PET groups (all *p* < 0.001, **Fig. 1B**). When stratifying the sample by Aβ-PET status, the results were similar such that in both groups, plasma p-tau217 levels increased over time (Aβ-positive: β = 0.10 to 0.12, *p* < 0.001; Aβ-negative: β = 0.07 to 0.09, *p* < 0.001, **Fig. 1A**). As a complementary measure to increase interpretability, we calculated the annual percent change for each assay. C2N p-tau217 had the highest annual percent change (12.31%), Fujirebio (10.65%), C2N %p-tau 217 (8.89%), Janssen (7.05%) and AlZpath (7.78%) in the whole group. The order of assays from highest to lowest annual percent change was similar in Aβ-positive (13.78% to 8.42%) and negative (11.48% to 6.27%) groups, although the magnitude was higher in the positive group. All details are reported in **Table S1**. Across plasma Aβ42/40 assays, only Aβ42/40 concentrations measured with Quanterix decreased significantly over time (β = -0.05, *p* < 0.001) in the whole group. There was no interaction between plasma Aβ42/40 levels over time and Aβ status (**Fig. 1B**). When stratifying the sample by Aβ-PET status, only a small decrease of Aβ42/40 over time was seen in the Aβ-negative group on the C2N and Quanterix assays (β = -0.02, *p* = 0.02 and β = -0.06, *p* < 0.001 respectively; **Fig. 1A**). The annual percent change was -0.10% to 0.85% in the full sample (**Table S1**).

**Figure 1.**
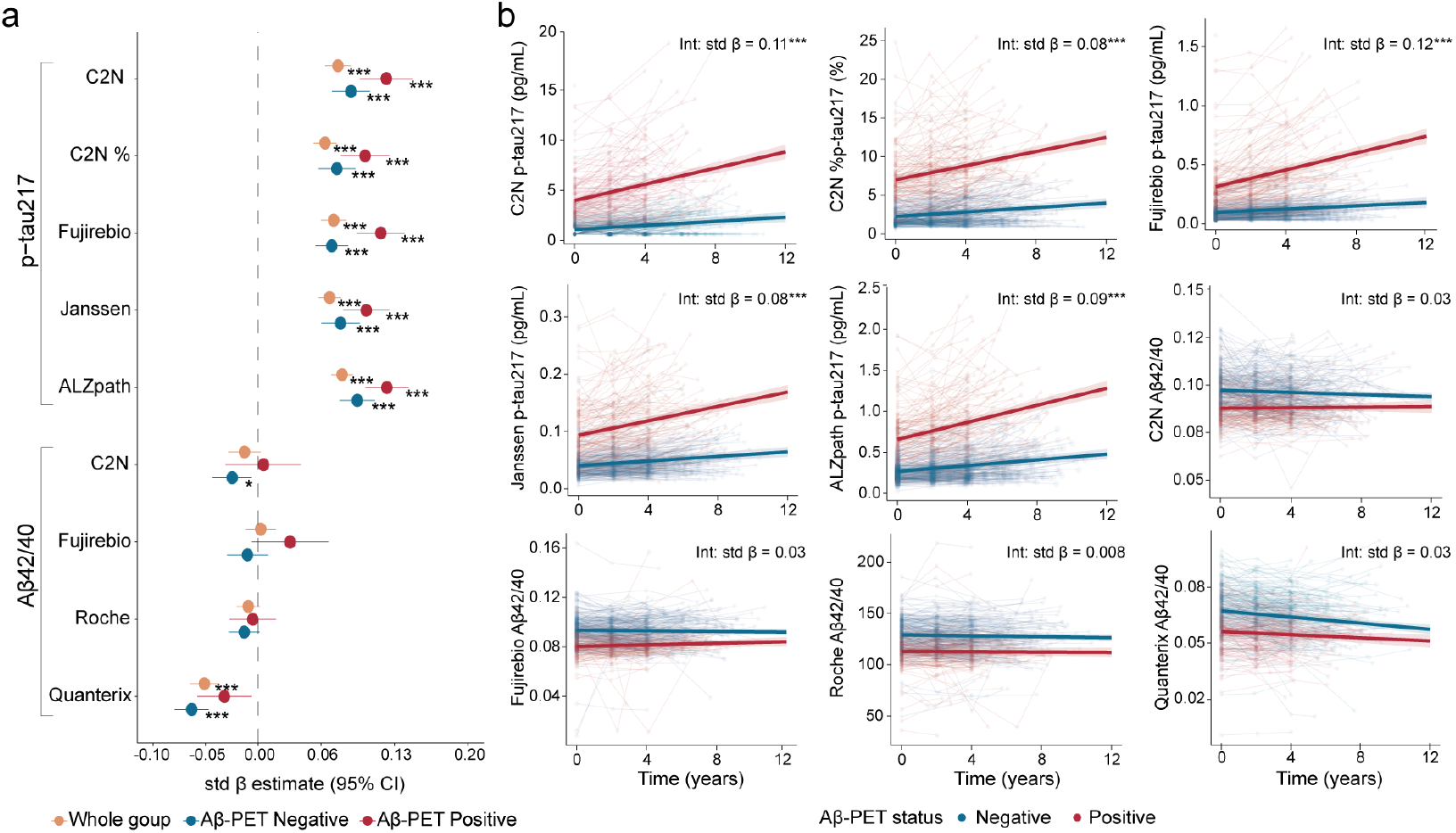
Longitudinal plasma biomarker trajectories across plasma p-tau217 and Aβ42/40 assays. **A)** Standardized regression coefficients (std β) and 95% confidence interval (CI) from linear-mixed effect models assessing plasma levels over time (plasma ∼ time) across the whole group (orange) and across the Aβ-negative (blue) and positive (red) groups. **B)**. Different plasma biomarkers over time with the interaction between time and Aβ status at baseline. The *x* axis shows time from first plasma biomarker sample. The std β and p-values of the interaction time × Aβ status are reported. All models were adjusted for age at first plasma timepoint, years of education and sex. \**p* < 0.05, \*\**p* < 0.005, \*\*\**p* < 0.001 after FDR correction for multiple comparisons across assays for each biomarker.

Among the other biomarkers measured, p-tau181, NfL and GFAP showed increased levels over time across the whole sample, as well as when stratified by Aβ-PET status (**Fig. S2**): Aβ-positive (β = 0.08 to 0.14, *p* < 0.001) and Aβ-negative groups (β = 0.05 to 0.09, *p* < 0.001). Overall, all assays showed similar effects. The annual percent change for these marker range between (4.65 % to 6.63%) in the full sample (**Table S1**)

### Concurrent changes in plasma biomarkers and Aβ-PET

Next, we assessed the relations between plasma markers and Aβ-PET Centiloids, comparing cross-sectional associations and longitudinal associations. Specifically, for the longitudinal associations, we investigated concurrent change in plasma biomarkers and Centiloids over the same period of up to 10 years. As a complementary analyses, we also evaluated if between baseline plasma levels were related to subsequent Centiloids change. We estimated individual biomarker slopes and compared these longitudinal associations to the baseline associations. In line with expected results, the cross-sectional associations showed that higher baseline p-tau217 concentrations was associated with higher Centiloid values in Aβ-positive participants (β = 0.44 to 0.64, adj R^2^ = 0.17 to 0.40, *p* < 0.001, **Fig. 2A, Table S2)**. We also observed significant but weaker associations in the Aβ-negative group (β = 0.18 to 0.26, adj R^2^ = 0.06 to 0.10, *p* < 0.01, **Fig. 2A**). The longitudinal results were different. In the Aβ-positive group, on all assays, p-tau217 rate of change was not associated with further Centiloid change (adj R^2^ = 0.03 to 0.05, *p >* 0.05, **Fig. 2B)**. In the Aβ-negative group, and consistent across all assays, higher p-tau217 change was associated with Centiloid change (β = 0.42 to 0.54, adj R^2^ = 0.14 to 0.28, *p* < 0.001, **Fig. 2B**). Such results were recapitulated across the whole group, where quadratic relationships were observed between concurrent p-tau217 and Centiloid changes, with a plateauing at the highest rates of change (adj R^2^ ranging from 0.49 [C2N] to 0.41 [ALZpath], **Fig. 2C**). All comparisons between linear or quadratic fit are reported in **Table S3**.

**Figure 2.**
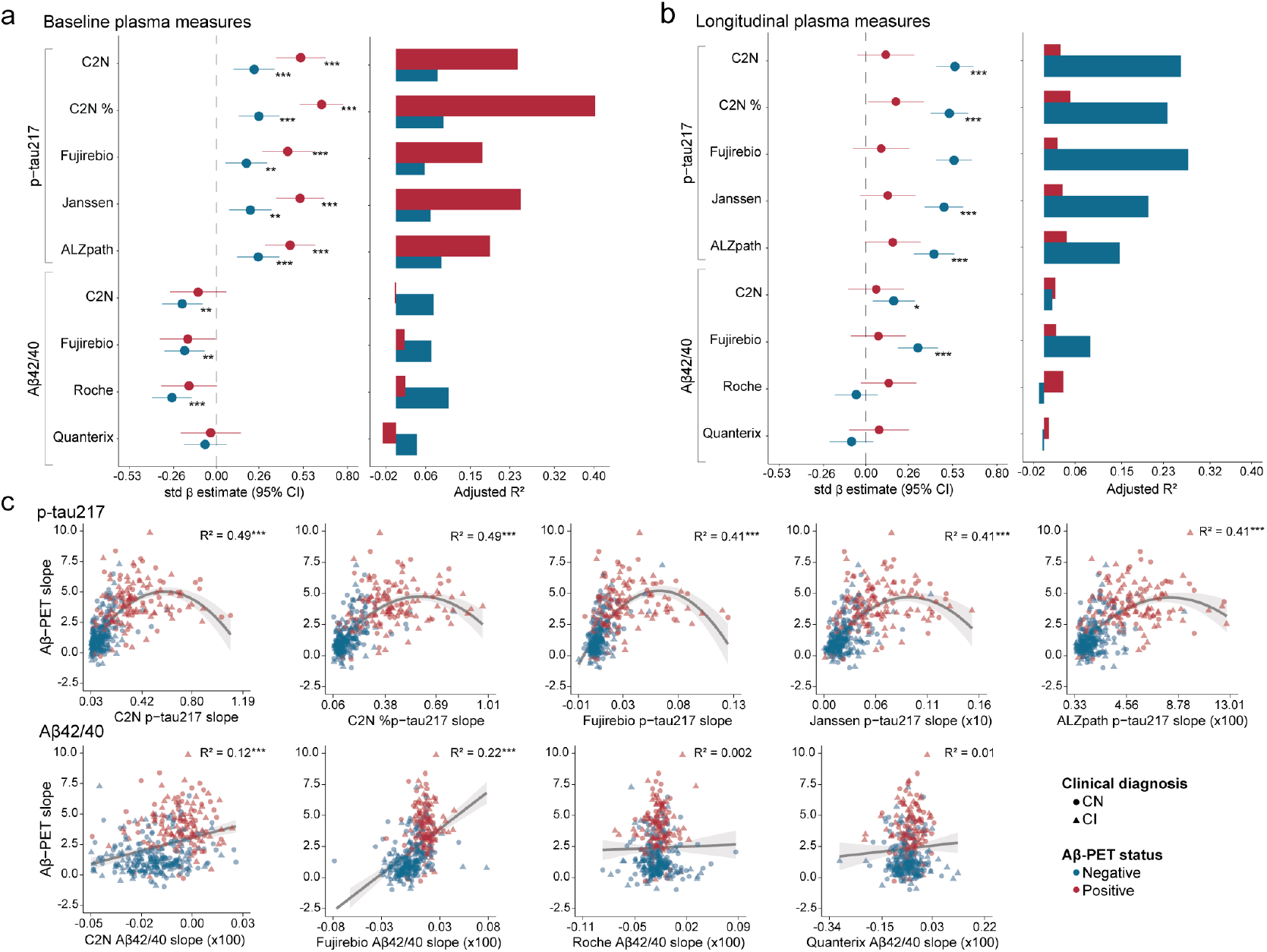
Association between longitudinal plasma biomarkers changes across plasma p-tau217 and Aβ42/40 with Aβ-PET Centiloids accumulation over time. **A)** Forest plots (left) of std β and 95% CIs and bar plots (right) of adjusted R^2^ for the Aβ-PET negative group (blue) and the Aβ-PET positive group (red), derived from linear regression models of baseline plasma biomarkers association with baseline Centiloids. **B)** Similar plots as in A derived from linear regression models of plasma biomarkers change and Centiloid change. **C)** Scatterplots of plasma levels rate of change (x-axis) and the Centiloid rate of change (y-axis) in the whole group with quadratic and linear regression applied where appropriate. All models were adjusted for age at first plasma timepoint, years of education and sex. \**p* < 0.05, \*\**p* < 0.005, \*\*\**p* < 0.001 after FDR correction for multiple comparisons across assays for each biomarker.

In cross-sectional associations with Aβ42/40 assays, lower levels at baseline were associated with higher Centiloid values in Aβ-negative individuals in all but one (Quanterix) assay (β = -0.27 to - 0.19, adj R^2^ = 0.07 to 0.11, *p* < 0.01, **Fig. 2A, Table S2**). However, no associations were found in the Aβ-positive group. Over time, there were almost no associations between Aβ42/40 rate of change and Centiloids change when stratified by Aβ-PET status. One exception was a positive association between concurrent Aβ42/40 and Centiloid change with the C2N and Fujeribio assays in the Aβ-negative group and when assessed in the full sample. We repeated similar associations with Aβ42 alone to clarify such results, where we found that reduced plasma Aβ42 change was associated with Centiloids change consistently across all assays (**Fig. S3 in Supplement**). The unexpected results with the C2N and Fujirebio Aβ42/40 ratios are likely due to the small rate of change and a more bimodal distribution between Aβ-groups in these assays.

Amid the recent US Food and Drug Administration approval of plasma p-tau217/Aβ42 from Fujirebio Lumipulse^32^, we performed similar analyses with this ratio, and found that the results were consistent, but with lower magnitude, with those seen using changes in p-tau217 alone (**Table S4)**.

In supplementary analyses, we investigated the trajectories of plasma p-tau181, NfL and GFAP in relation to Aβ-PET rate of change (**Fig. S4**). In the whole group, a linear association was seen between higher plasma changes and Centiloids change, with the highest associations for p-tau181 and GFAP (adj R^2^ = 0.19-0.12), and the lowest ones with NfL assays (adj R^2^ = 0.03-0.05, **Table S3**). Associations were also seen in the Aβ-negative group for p-tau181 and GFAP, where plasma changes were associated with higher Centiloid change (β = 0.23 to 0.33, adj R^2^ = 0.05 to 0.11, *p* < 0.05, **Fig. S4**).

As a comparison, we also examined how well baseline plasma biomarker levels related to subsequent Centiloid change. Associations were only seen on the whole group and in the Aβ-negative group (**Fig. S5, Table S5**). In the whole group, only baseline p-tau217 levels were related to Centiloids change but explained lower variance (adj R^2^ from 0.37 to 0.25) than the longitudinal results (adj R^2^ from 0.49 to 0.41). In the Aβ-negative group, higher p-tau217 (adj R^2^ = 0.02 to 0.08, *p* < 0.01) and lower Aβ42/40 (adj R^2^ = 0.02 to 0.11, *p* < 0.01) levels at baseline were associated with subsequent Centiloids change, but the magnitude was lower than the longitudinal associations (**Fig. S5)**.

### Relations between plasma biomarkers and subsequent Aβ positivity

Seeing the consistent associations between plasma levels change and Centiloids change in the Aβ-negative group, we investigated the ability of plasma markers to predict progression to Aβ positivity with Cox-proportional hazard ratio models. Among the 249 Aβ-negative participants at baseline, 52 (20.88%) progressed to Aβ-PET positivity over a mean (SD) of 3.86 (2.09) years. Across all p-tau217 assays, higher levels were associated with progression to Aβ-positivity (HR from 1.59 to 2.04, all *p* < 0.001, **Fig. 3A-B**). However, plasma rate of change was a highest predictor of future Aβ-PET positivity across all assays (HR range from 1.90 to 2.69, all *p* < 0.001, **Fig. 3A-B**), with p-tau217 C2N assay showing the highest association (**Fig. 3C**)

**Figure 3.**
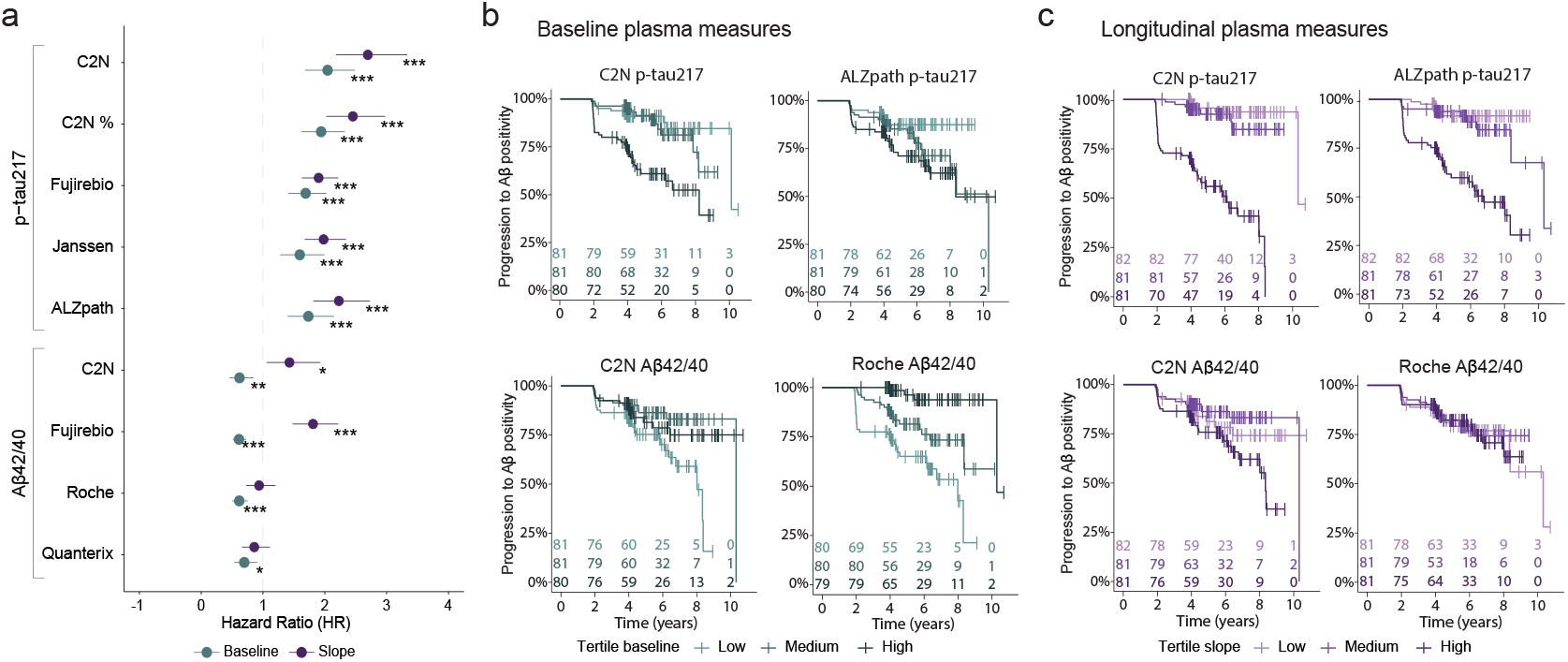
Progression to Aβ-PET positivity in baseline Aβ-PET negative participants in plasma p-tau217 and Aβ42/40 biomarkers. **A)** Forest plots showing the HRs and 95% CIs derived from survival analyses using baseline plasma biomarkers and slopes. **B)** Survival curves for progression to Aβ-PET positivity across different plasma p-tau217 and Aβ42/40 at baseline. **C)** Survival curves for progression to Aβ-PET positivity across different plasma p-tau217 and Aβ42/40 slopes. All models were adjusted for age at first plasma timepoint, years of education and sex. Continuous values were used for statistical analyses, and plasma biomarkers were divided into tertiles for visualization. N = 52 participants progressed to Aβ-PET positive over the course of follow-ups, N = 197 participants remained Aβ-PET negative. \**p* < 0.05, \*\**p* < 0.005, \*\*\**p* < 0.001 after FDR correction for multiple comparisons across assays for each biomarker.

Among all plasma Aβ42/40 assays, lower concentration at baseline was associated with an increased risk of progression to Aβ-positivity (HR from 0.61 to 0.70 [1.42 to 1.64 if reversed to go in the same direction as p-tau217], *p* < 0.01, **Fig. 3A-B**). There were no associations with plasma rate of change, except an unexpected positive association with C2N and Fujirebio Lumipulse assays (HR = 1.43, *p* = 0.04; HR = 1.81, *p* < 0.001 respectively; **Fig. 3A-C**), as seen in the previous analyses. When instead using Aβ42 rate of change alone, we found that, as expected, reduced Aβ42 rate of change was associated with future Aβ-PET positivity across all assays (HR from 0.55 to 0.64, *p* < 0.001, **Table S6**).

Finally, higher plasma p-tau181 and GFAP rate of change, but not their baseline levels were associated with progression to Aβ positivity (HR = 1.35 to 1.75, *p* < 0.01, **Table S6**). No associations were seen with NfL. For visualisation, plasma rate of change on all assays across the three groups, i.e. individuals who remained Aβ-PET negative, who progressed to Aβ-PET positivity and who were Aβ-positive are shown in **Fig. S6**.

### Associations between plasma biomarkers and longitudinal cognition

Lastly, we evaluated whether change in plasma biomarkers was related to cognitive decline, by calculating change on the Alzheimer’s Disease Assessment Scale (ADAS13, where higher ADAS13 scores represents worse performance). Results were also corroborated with Mini-Mental State Examination (MMSE; **Fig. S7**). First, using cross-sectional associations, we found that all p-tau217 assays were associated with worse cognition at baseline, only in Aβ-positive participants (β = 0.28 to 0.43, adj R^2^ = 0.08 to 0.19, *p* < 0.001; **Fig. 4A**). Longitudinal plasma measures showed more significant associations with cognitive decline, all of which followed a linear pattern except for %C2N p-tau217 that followed a quadratic fit. In Aβ-positive individuals, higher plasma p-tau217 rate of change was associated with greater worsening in ADAS13 scores (β = 0.50 to 0.63, adj R^2^ = 0.21 to 0.36; *p* < 0.001; **Fig. 4B**). Similar results were observed in the whole sample (**Fig. 4C**). The association between plasma and cognitive rate of change was also observed in the Aβ-negative group with a smaller magnitude (β = 0.11 to 0.19, adj R^2^ = 0.07 to 0.10; *p* < 0.001; **Fig. 4B**). Only minimal associations were seen with Aβ42/40 assays (**Fig. 4**). In comparison, associations between baseline Centiloids and baseline cognition explained 0.14 variance and the association between Centiloids and cognitive slopes explained 0.12 variance (**Fig. S8**).

**Figure 4.**
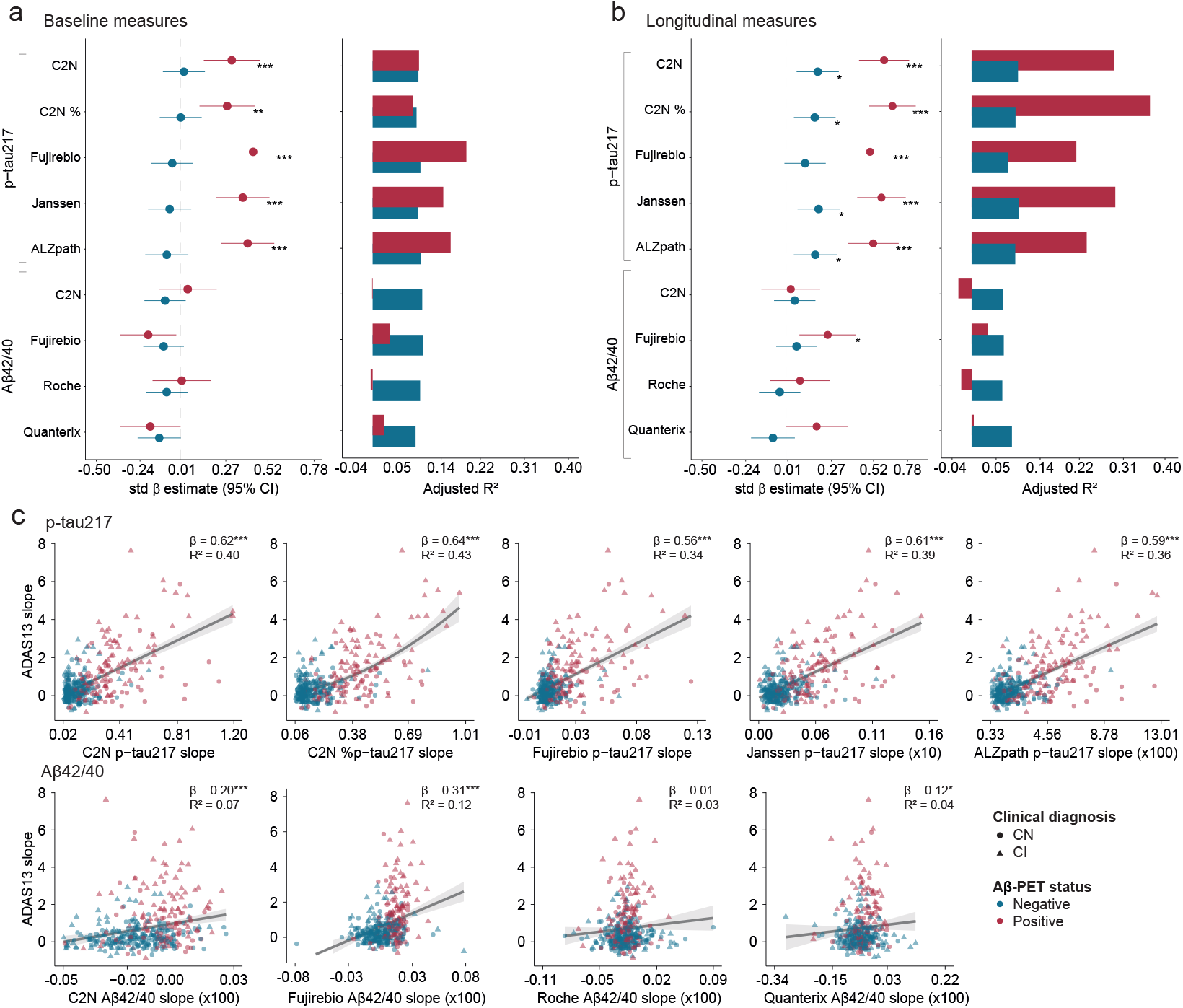
Association between longitudinal plasma biomarkers and ADAS13 rate of change. **A)** Forest plots (left) of std β and 95% CIs and bar plots (right) of adjusted R^2^ for the Aβ-PET negative group (blue) and the Aβ-PET positive group (red), derived from linear regression models of baseline plasma biomarkers association with baseline ADAS13 score. **B)** Similar plots as in A derived from linear regression models of plasma biomarkers change and change in ADAS13. **C)** Scatterplots showing the rate of change in plasma levels (x-axis) and ADAS13 (y-axis) in the whole group with quadratic (% C2N p-tau217) and linear regression applied where appropriate. The std β, adj R^2^ and p-values from linear model for %C2N p-tau217 is reported. All models were adjusted for age at first plasma timepoint, years of education and sex. ADAS13: Alzheimer’s Disease Assessment Scale-Cognitive Subscale 13. \**p* < 0.05, \*\**p* < 0.005, \*\*\**p* < 0.001 after FDR correction for multiple comparisons across assays for each biomarker.

For p-tau181, GFAP and NfL, associations between plasma change and cognitive decline were found in the whole sample and in the Aβ-positive groups, with higher variance explained than for cross-sectional associations (**Table S7**, adj R^2^ = 0.09-0.17 for longitudinal vs 0.07-0.12).

## DISCUSSION

In this longitudinal head-to-head plasma study spanning over a decade of data, we assessed the trajectories of five plasma biomarkers measured across fourteen assays, their association with concurrent changes in Aβ deposition and with cognitive decline. We found that all plasma markers increased over time, except for the Aβ42/40 assays that was mainly stable over the 10 years window. When stratified by baseline Aβ-PET status, rate of change in plasma p-tau217 increased with accumulating insoluble Aβ-PET burden in individuals who had not yet reached Aβ-PET positivity. On the other hand, in Aβ-positive individuals, the rate of change of plasma p-tau217 was no longer associated with a concurrent Aβ-PET accumulation. Further, among Aβ-negative individuals, changes in plasma p-tau217 strongly predicted future progression to Aβ-PET positivity. These results support the hypothesis that plasma p-tau217 levels increase as Aβ begins to accumulate, years before individuals reach a positive threshold for Aβ-PET.^33, 34^ This early change may explain the disconnection between longitudinal measures of plasma and Aβ pathology when individuals surpassed the Aβ-PET positivity threshold (CL = 20). Regarding cognition, the rate of change in plasma p-tau217 was associated with worsening cognitive performance over time, especially in the Aβ-PET positive group. While change in plasma p-tau217 might no longer reflect additional accumulation of insoluble Aβ pathology in individuals with significant Aβ plaques, it captures clinically meaningful decline at this later disease stage. Taken together, these findings indicate strong potential of the use of plasma biomarkers changes, especially p-tau217, as surrogate markers of accumulating pathology early in the disease course and as predictors of cognitive decline later in the disease course.

A key finding of this study is the meaningful change observed in plasma p-tau biomarkers, particularly p-tau217, which increases as people progress toward Aβ-PET positivity reflecting their performance in predicting pathology.^26, 35^ These observations were consistent across platforms, regardless of the assay, and align with recent meta-analyses and cross-sectional studies showing that all plasma p-tau217 assays exhibit high performance in detecting Aβ positivity.^9, 17, 36^ Such results are important as they underscore the potential of p-tau217 change to track earliest signs of accumulating Aβ pathology before someone is positive on a PET scan, which has important implications for monitoring patients in clinical settings as well as those under anti-amyloid therapies.^37^ Across the whole group, changes in plasma p-tau217 showed non-linear association with concurrent change in Aβ, suggesting a plateauing of p-tau217 levels in relation to Aβ change at higher levels of Aβ pathology. This observation is in line with a recent cross-sectional study that showed non-linear trend in %p-tau217 with higher levels of Aβ-PET, followed by a plateau after exceeding the Aβ-PET threshold suggesting the role of p-tau217 in identifying early AD pathological change.^38^ Relatedly and importantly, plasma p-tau217 rates of change also predicted subsequent Aβ positivity over on average 3-4 years later, with higher associations than the baseline p-tau217 levels. This was also true for plasma p-tau181 and GFAP, in which only their rate of change was associated with subsequent Aβ positivity.^39^

While the changes in plasma p-tau217 markers seem highly sensitive in the early pathological phases, they may have limited utility for tracking accumulating Aβ pathology beyond the point of PET-confirmed positivity. In the Aβ-positive individuals, changes across all plasma p-tau217 assays related to cognitive decline, with some assays explaining more than twice as high variance compared to the cross-sectional associations. These results align with the fact that p-tau markers reflect both Aβ and tau pathologies, as demonstrated in neuropathological studies.^5^ Although tau-PET was not available in our study, elevated p-tau change in the Aβ-positive group may still reflect ongoing tau aggregation, which in turn increases the risk of accelerated cognitive decline.^40^ Collectively, our findings highlight the prognostic potential of plasma p-tau217 change. Among other markers tested, plasma NfL slopes were not associated with concurrent Aβ accumulation, but with cognitive decline, suggesting their added value for changes occurring in later disease progression.^41^

As for plasma Aβ42/40, we observed almost no changes across assays over the 10 years follow-up, and no association were seen with accumulating pathology or cognitive decline. Rather, cross-sectional levels of Aβ42/40 were associated with Aβ pathology or cognition. Unexpectedly, two assays, C2N and Fujirebio Lumipulse, out of the four available exhibited increased rate of change in plasma Aβ42/40 with greater Aβ accumulation while associations with change in Aβ42 alone showed the expected negative association. This discordance may be attributed to the narrow range of longitudinal changes in plasma Aβ42/40 levels, and affecting the distribution between Aβ-positive and negative participants. Such biomarkers have also been reported as the ones changing the earliest before the pathology is detectable on PET^26, 42^, and given our results we speculate that such changes might occur very on, prior to the time range of the current study.Taken together, given that either no associations or unexpected associations were seen with Aβ42/40 change and any outcomes of interest in this study, we conclude that that longitudinal change in this biomarker has limited value compared to its cross-sectional measurements.

The main strength of the study is the longitudinal head-to-head design of plasma measures across 15 assays, Aβ-PET and clinical data spanning over a decade. This has allowed to compare cross-sectional and longitudinal associations, highlighting key results where plasma rates of change are very promising. However, there are limitations to consider. Tau-PET was not included in our study given the limited number of ADNI participants with available longitudinal tau-PET, and we hypothesize that in Aβ-positive individuals, plasma rates of change might better relate to accumulating tau tangles than Aβ load.^27^ In addition, the ADNI cohort consists primarily of White and highly educated individuals. Future studies should aim to replicate our findings in more diverse, real-world populations.

In conclusion, we showed that changes in plasma biomarkers, particularly p-tau217, displayed longitudinal changes associated to different factors along the AD continuum. In early stages of the disease, higher changes in plasma p-tau217 reflected accumulating Aβ pathology, highlighting the potential of p-tau217 as a valuable predictive marker in the context of emerging anti-amyloid therapies. On the other hand, in people with elevated Aβ, higher changes in plasma markers predicted faster cognitive decline rather than Aβ accumulation suggesting the utility of these markers in monitoring disease progression in clinical settings and trials. Our results highlight the dual utility of longitudinal p-tau217 levels as both a surrogate marker of pathology early on and a tool for monitoring cognition later in the disease process.

## MATERIALS AND METHODS

### Participants

We included 394 participants from ADNI, a non-randomized multi-site study launched in 2003 as a public-private partnership. The primary goal of ADNI has been to test whether serial MRI, PET, other biological markers, and clinical and neuropsychological assessment can be combined to measure the progression of MCI and early AD. For up-to-date information, see www.adni-info.org.

To be included in the study, participants had to be between 55-90 years old at baseline, to score < 6 on Geriatric Depression Scale, to have a study partner with 10+ hours per week who could accompany them and no diseases that could preclude their enrollment^43^. Clinical diagnosis was evaluated among neurologists and dementia experts in addition to neuropsychological assessment blinded to the biomarkers results. CU individuals had Clinical Dementia Rating Scale (CDR) of 0 and Mini Mental state Exam between 24 and 30, participants with MCI had a CDR of 0.5 and MMSE between 24 and 30, participants with AD dementia had CDR of 0.5 or 1 and MMSE between 20 and 24. For the purpose of the study, we included participants included in the FNIH consortium, such that they had plasma and Aβ-PET available at an average of three time points. The ADNI study was approved by the Institutional Review Board of each participating site and written informed consent was obtained from all participants.

### Biomarker measurements Plasma biomarker assays

Plasma samples were collected according to ADNI protocol. Plasma samples were measured blinded to the participant’s information. The following analytes, Aβ_40_, Aβ_42_, p-tau217, p-tau181, NfL, and GFAP, were quantified across 14 assays.^44^ As such, Aβ_40_, Aβ_42_, p-tau217, unphosphorylated tau217 concentrations were measured using the C2N PrecivityAD2 liquid chromatography tandem mass spectrometry (LC‐MS) based assay at C2N Diagnostics laboratory in St. Louis, USA. %p‐tau217 was calculated as the ratio of p-tau217 to unphosphorylated tau217 multiplied by 100. Plasma Aβ_40_, Aβ_42_, p-tau217 were also measured using Fujirebio Lumipulse commercial kits on a Fujirebio Lumipulse G1200 analyzer at the Indiana University National Centralized Repository for Alzheimer’s Disease and Related Dementias Biomarker Assay Laboratory. Plasma Aβ_40_, Aβ_42,_ p-tau181, NfL and GFAP were measured using the Roche NeuroToolKit on a Cobas e801 analyzer (Roche Diagnostics International Ltd, Switzerland) at the University of Gothenburg; and using Quanterix Neurology 4‐Plex E (N4PE) assays for Aβ42, Aβ40, p‐tau181 (v2.1), NfL, and GFAP on Quanterix Simoa‐HD‐X analyzer at the Quanterix Accelerator Laboratory. P-tau217 were also analyzed using Quanterix, ALZpath and Janssen assay in the same laboratory. All measurements were made in singlicate except for Quanterix based assays, that were run in duplicates. Further details have been described in the main paper describing cross-sectional results on this data from the FNIH Consortium.^44^ A total of 27 participants had missing values in at least one timepoint across all fourteen assays. A maximum of 55 values were removed, with Quanterix’s Neurology 4-Plex p-tau181, Aβ42/40, GFAP, and NfL (all details per assay are reported in **Table S8**). Twelve participants have missing timepoints across all markers. For each assay, values above or below 5 standard deviations from the mean across all available timepoints were considered outliers, with a with a maximum of 6 outliers identified in Fujrebio p-tau217 and in Quanterix p-tau181 (all details per assay are reported in **Table S8**).

### PET processing

We used Aβ-PET data processed by the ADNI PET Core. Aβ-PET scans were acquired using two tracers ([^18^F] florbetaben with acquisition 90-110 minutes post-injection and [^18^F] florbetapir with acquisition 50-70 minutes post-injection). In brief, frames were aligned, averaged and smoothed to 8 mm^3^ and registered to closest T1 MPRAGE image that had been processed with FreeSurfer. Details on PET acquisition and processing have been described elsewhere (https://adni.loni.usc.edu/documents/pet). Scans that failed quality control (n = 4) were excluded from the analyses. Global amyloid-PET standardized uptake value ratio (SUVR) was defined as the weighted average of tracer retention in frontal, lateral parietal, anterior/posterior cingulate and lateral temporal cortices using cerebellum as a reference region. SUVR values were converted to the Centiloid scale using Klunk et al method^45^. The threshold for Aβ-PET positivity was set as > 20 Centiloid.^44^ We included all Aβ-PET scans available during the same period as all plasma measurements, resulting in 3 or 4 PET scans per participant. All PET scans retained were acquired within one year of each plasma measures, except for five participants whose last scans were excluded due to a two-year gap between plasma and PET acquisition. This rendered two scans for those participants (3 participants are Aβ-PET positive, 2 participants are Aβ-PET negative).

### Neuropsychological evaluations

We utilized the ADAS-Cog 13, a composite cognitive assessment that includes measures of attention and concentration, executive function and planning, verbal and nonverbal memory, praxis, delayed word recall, and number cancellation or maze tasks. Scores range from 0 to 85, with higher scores indicating greater cognitive impairment.^46^ In supplementary analyses, we also examined the associations between longitudinal plasma biomarkers and cognitive performance as measured by the MMSE (**see Supplement**).

### Statistical analyses

Participants characteristics between Aβ-PET negative and positive individuals were compared using Mann-Whitney U test and chi-squared test for categorical variables. Spearman correlations were used to evaluate the relationships between continuous baseline and longitudinal plasma, Aβ-PET and cognitive measures. At the group level, to evaluate how plasma concentrations change over time, we report the effect of time from linear mixed effect models (LME) with random slope and intercept. As a complementary analysis, we calculated the annualized percent change for each plasma biomarker using the baseline and last available follow-up measurement for each participant. Such analyses were done across the entire sample and for the Aβ-PET positive and negative groups separately. At the individual level, as a measure of individual rate of change in plasma and Aβ-PET, we extracted the individual slope from LME using longitudinal plasma concentrations or Aβ-PET Centiloid as an outcome and time (years from first plasma time point) as a predictor with random slope and intercept. We then conducted linear regressions to assess the relations between concurrent plasma and Centiloid rates of change as well as relations between baseline plasma concentrations and baseline Centiloid. We selected the first available plasma measure and closest Aβ-PET scan for baseline analyses. Such analyses were conducted in the whole sample, as well as in Aβ-PET positive and negative groups separately. For analyses done on the whole group, to determine whether the association from regression models had a linear or quadratic fit, we compared the Akaike Information Criterion (AIC) and the Bayesian Information Criterion (BIC) from both models. In cases where ΔAIC and ΔBIC > 10 (defined as the difference between linear and quadratic models), we considered that a quadratic fit was better. In complementary analyses, we conducted similar associations as described above, looking at associations between baseline plasma concentrations and Centiloid rate of change. In a next set of analyses, we assessed progression to Aβ-PET positivity. We stratified Aβ-PET negative participants into those who remained negative across all time points vs those who progressed to Aβ-PET positivity. Two participants were excluded from the analyses as they reverted to negative at all follow-up scans. We used cox proportional-hazards models to assess if baseline plasma concentrations and/or plasma rate of change were associated with subsequent Aβ-PET positivity. For individuals who became positive, we calculated the time difference between the date of the first available positive Aβ-PET scan and the baseline scan, while the time difference between the last available Aβ-PET scan and the baseline scan for those who remained negative. For cognitive outcomes, we repeated the initial set of analyses using ADAS-Cog13 and MMSE scores as outcomes instead of Aβ-PET pathology. Across all analyses, all models were adjusted for age at first plasma timepoint, years of education and sex. Models were adjusted for multiple comparisons by the number of assays for each biomarker of interest (Aβ42/40, p-tau217, p-tau181, NfL, GFAP) using FDR method. All analyses were carried out in R (4.4.1).

## ACKNOWLEDGEMENT

The Foundation for the National Institutes of Health (FNIH) Biomarkers Consortium project was made possible through the combined scientific and financial support of industry, academia, government, and patient advocacy partners. We thank the following project team members for their valuable contributions: Anthony Bannon (AbbVie), Lei Du-Cuny (AbbVie), Yulia Mordashova (AbbVie), William Potter, Maria Quinton (AbbVie), Christopher Weber (Alzheimer’s Association), Emily Meyers (Alzheimer’s Association), Patricia Saletti (Alzheimer’s Drug Discovery Foundation), Kyle Ferber (Biogen), Carrie Rubel (Biogen), Erin Rosenbaugh (FNIH), Jeff Dage (Indiana University), Ziad Saad (Johnson and Johnson Innovative Medicine), Gallen Triana-Baltzer (Johnson and Johnson Innovative Medicine), Hartmuth Kolb (formerly with Johnson and Johnson Innovative Medicine), John Hsiao (NIA), Michael Baratta (Takeda), Janaky Coomaraswamy (Takeda), Iwona Dobler (Takeda), Dave Raunig (Takeda), Henrik Zetterberg (University of Gothenburg), Leslie Shaw (University of Pennsylvania), Suzanne Schindler (Washington University in St. Louis). The authors recognize C_2_N Diagnostics, Fujirebio Diagnostics with the Indiana University National Centralized Repository for Alzheimer’s Disease and Related Dementias Biomarker Assay Laboratory (NCRAD-BAL), Quanterix, and Roche Diagnostics with the University of Gothenburg for performing the plasma biomarker analysis in this study. We are thankful to Drs Nicholas Ashton and Henrik Zetterberg and the University of Gothenburg for generating the plasma endogenous QC pools confirmed high or low for pTau217. COBAS and ELECSYS are trademarks of Roche. All other product names and trademarks are the property of their respective owners. The NeuroToolKit is a panel of exploratory prototype assays designed to robustly evaluate biomarkers associated with key pathologic events characteristic of AD and other neurological disorders, used for research purposes only and not approved for clinical use (Roche Diagnostics International Ltd, Rotkreuz, Switzerland). We would like to acknowledge the invaluable contributions of ADNI participants, along with the ADNI investigators and research administration team, whose efforts made these data openly available.

## AUTHOR CONTRIBUTIONS

Y.Y., A.P.B. and S.V. designed the study and drafted the manuscript. A.P.B. led the investigation and S.V. contributed to supervision. Y.Y. and A.P.B. carried out data curation and statistical analyses of the manuscript. T.Q. provided statistical support. C.P. helped in figure preparation. All authors contributed to the interpretation of the results, reviewed and approved the final version of the manuscript.

## FUNDING

Data collection and sharing for the Alzheimer’s Disease Neuroimaging Initiative (ADNI) is funded by the National Institute on Aging (National Institutes of Health Grant U19AG024904). The grantee organization is the Northern California Institute for Research and Education. In the past, ADNI has also received funding from the National Institute of Biomedical Imaging and Bioengineering, the Canadian Institutes of Health Research, and private sector contributions through the Foundation for the National Institutes of Health (FNIH) including generous contributions from the following: AbbVie, Alzheimer’s Association; Alzheimer’s Drug Discovery Foundation; Araclon Biotech; BioClinica, Inc.; Biogen; Bristol-Myers Squibb Company; CereSpir, Inc.; Cogstate; Eisai Inc.; Elan Pharmaceuticals, Inc.; Eli Lilly and Company; EuroImmun; F. Hoffmann-La Roche Ltd and its affiliated company Genentech, Inc.; Fujirebio; GE Healthcare; IXICO Ltd.; Janssen Alzheimer Immunotherapy Research & Development, LLC.; Johnson & Johnson Pharmaceutical Research & Development LLC.; Lumosity; Lundbeck; Merck & Co., Inc.; Meso Scale Diagnostics, LLC.; NeuroRx Research; Neurotrack Technologies; Novartis Pharmaceuticals Corporation; Pfizer Inc.; Piramal Imaging; Servier; Takeda Pharmaceutical Company; and Transition Therapeutics. Funding partners of the FNIH Biomarkers Consortium include AbbVie Inc., Alzheimer’s Association®, Diagnostics Accelerator at the Alzheimer’s Drug Discovery Foundation, Biogen, Janssen Research & Development, LLC, and Takeda Pharmaceutical Company Limited. Private-sector funding for the study was managed by the Foundation for the National Institutes of Health. The NCRAD-BAL is supported by a cooperative agreement grant (U24 AG021886) awarded to NCRAD by the National Institute on Aging. Y.Y. received support from Alzheimer’s Society of Canada foundation and Fonds de recherche du Québec-Santé. S.V. is supported by the Alzheimer Society of Canada, the Alzheimer’s Association, the Tier-1 Canada Research Chair in Early Detection of Alzheimer’s Disease and the Canadian Institutes of Health Research (CIHR, 178385, 162091, 148963). A.P.B. received funding from FRQ (361227) and the Fondation de l’Institut de gériatrie de Montréal. GS received funding from the European Union’s Horizon 2020 Research and Innovation Program under Marie Sklodowska-Curie action grant agreement number 101061836, an Alzheimer’s Association Research Fellowship (AARF-22-972612), the Brightfocus Foundation (A2024007F), the Alzheimerfonden (AF-980942, AF-994514, AF-1012218), Greta och Johan Kocks research grants and travel grants from the Strategic Research Area MultiPark (Multidisciplinary Research in Parkinson’s Disease) at Lund University, and from the ADDF at BBRC.

## DATA AVAILABILITY

The data used in this study were obtained from the publicly available ADNI dataset, accessible through the ADNI database (adni.loni.usc.edu). A complete list of ADNI investigator can be found at:http://adni.loni.usc.edu/wp-content/uploads/how_to_apply/ADNI_Acknowledgement_List.pdf.

## CODE AVAILABILITY

The code used in the preparation of this manuscript is available upon request.

## ETHICS DECLARATIONS COMPETING INTERESTS

GS has received speaker fees from Springer, GE Healthcare, Biogen, Esteve and Adium and advisory fees from Johnson&Johnson. The authors declare no competing interests

## Notes

### Author Declarations

The ADNI study was approved by the Institutional Review Boards of all participating institutions. Written informed consent was obtained for all participants or their authorized representative.

## References

1. Schöll M, Verberk IMW, del Campo M, et al. Challenges in the practical implementation of blood biomarkers for Alzheimer’s disease. The Lancet Healthy Longevity. 2024;5(10)doi:10.1016/j.lanhl.2024.07.013

2. Palmqvist S, Whitson HE, Allen LA, et al. Alzheimer’s Association Clinical Practice Guideline on the use of blood-based biomarkers in the diagnostic workup of suspected Alzheimer’s disease within specialized care settings. Alzheimer’s & Dementia. 2025/07/01 2025;21(7):e70535. doi:10.1002/alz.70535

3. Schindler SE, Galasko D, Pereira AC, et al. Acceptable performance of blood biomarker tests of amyloid pathology — recommendations from the Global CEO Initiative on Alzheimer’s Disease. Nature Reviews Neurology. 2024/07/01 2024;20(7):426–439. doi:10.1038/s41582-024-00977-5

4. Barthélemy NR, Salvadó G, Schindler SE, et al. Highly accurate blood test for Alzheimer’s disease is similar or superior to clinical cerebrospinal fluid tests. Nature medicine. 2024;30(4):1085–1095. doi:10.1038/s41591-024-02869-z

5. Salvadó G, Ossenkoppele R, Ashton NJ, et al. Specific associations between plasma biomarkers and postmortem amyloid plaque and tau tangle loads. EMBO Molecular Medicine. n/a(n/a):e2463. doi:10.15252/emmm.202217123

6. Palmqvist S, Warmenhoven N, Anastasi F, et al. Plasma phospho-tau217 for Alzheimer’s disease diagnosis in primary and secondary care using a fully automated platform. Nature Medicine. 2025/06/01 2025;31(6):2036–2043. doi:10.1038/s41591-025-03622-w

7. Mattsson-Carlgren N, Janelidze S, Bateman RJ, et al. Soluble P-tau217 reflects amyloid and tau pathology and mediates the association of amyloid with tau. EMBO Mol Med. Jun 7 2021;13(6):e14022. doi:10.15252/emmm.202114022

8. Ashton NJ, Keshavan A, Brum WS, et al. The Alzheimer’s Association Global Biomarker Standardization Consortium (GBSC) plasma phospho-tau Round Robin study. Alzheimer’s & Dementia. 2025/02/05 2025;n/a(n/a):e14508. doi:10.1002/alz.14508

9. Schindler SE, Petersen KK, Saef B, et al. Head-to-head comparison of leading blood tests for Alzheimer’s disease pathology. Alzheimer’s & dementia: the journal of the Alzheimer’s Association. 2024;20(11):8074–8096. doi:10.1002/alz.14315

10. Feizpour A, Doecke JD, Doré V, et al. Detection and staging of Alzheimer’s disease by plasma pTau217 on a high throughput immunoassay platform. eBioMedicine. 2024;109 doi:10.1016/j.ebiom.2024.105405

11. Ossenkoppele R, Salvadó G, Janelidze S, et al. Plasma p-tau217 and tau-PET predict future cognitive decline among cognitively unimpaired individuals: implications for clinical trials. Nature Aging. 2025/03/28 2025;doi:10.1038/s43587-025-00835-z

12. Yakoub Y, Gonzalez-Ortiz F, Ashton NJ, et al. Plasma p-tau217 identifies cognitively normal older adults who will develop cognitive impairment in a 10-year window. Alzheimer’s & Dementia. 2025;21(2):e14537. doi:10.1002/alz.14537

13. Ashton NJ, Janelidze S, Mattsson-Carlgren N, et al. Dinerential roles of Aβ42/40, p-tau231 and p-tau217 for Alzheimer’s trial selection and disease monitoring. Nature Medicine. 2022/12/01 2022;28(12):2555–2562. doi:10.1038/s41591-022-02074-w

14. Ashton NJ, Pascoal TA, Karikari TK, et al. Plasma p-tau231: a new biomarker for incipient Alzheimer’s disease pathology. Acta neuropathologica. 2021;141(5):709–724. doi:10.1007/s00401-021-02275-6

15. Meyer P-F, Ashton NJ, Karikari TK, et al. Plasma p-tau231, p-tau181, PET Biomarkers, and Cognitive Change in Older Adults. Annals of neurology. 2022;91(4):548–560. doi:10.1002/ana.26308

16. Suárez-Calvet M, Karikari TK, Ashton NJ, et al. Novel tau biomarkers phosphorylated at T181, T217 or T231 rise in the initial stages of the preclinical Alzheimer’s continuum when only subtle changes in Aβ pathology are detected. EMBO molecular medicine. 2020;12(12):n/a-n/a.

17. Janelidze S, Bali D, Ashton NJ, et al. Head-to-head comparison of 10 plasma phospho-tau assays in prodromal Alzheimer’s disease. Brain. Apr 19 2023;146(4):1592–1601. doi:10.1093/brain/awac333

18. Janelidze S, Teunissen CE, Zetterberg H, et al. Head-to-Head Comparison of 8 Plasma Amyloid-β 42/40 Assays in Alzheimer Disease. JAMA neurology. 2021;78(11):1375–1382. doi:10.1001/jamaneurol.2021.3180

19. Olsson B, Portelius E, Cullen NC, et al. Association of Cerebrospinal Fluid Neurofilament Light Protein Levels With Cognition in Patients With Dementia, Motor Neuron Disease, and Movement Disorders. JAMA Neurol. Mar 1 2019;76(3):318–325. doi:10.1001/jamaneurol.2018.3746

20. Ashton NJ, Janelidze S, Al Khleifat A, et al. A multicentre validation study of the diagnostic value of plasma neurofilament light. Nature Communications. 2021/06/07 2021;12(1):3400. doi:10.1038/s41467-021-23620-z

21. Chatterjee P, Doré V, Pedrini S, et al. Plasma Glial Fibrillary Acidic Protein Is Associated with 18F-SMBT-1 PET: Two Putative Astrocyte Reactivity Biomarkers for Alzheimer’s Disease. J Alzheimers Dis. 2023;92(2):615–628. doi:10.3233/jad-220908

22. Yang Z, Sreenivasan K, Toledano Strom EN, et al. Clinical and biological relevance of glial fibrillary acidic protein in Alzheimer’s disease. Alzheimers Res Ther. Nov 3 2023;15(1):190. doi:10.1186/s13195-023-01340-4

23. Bilgel M, An Y, Walker KA, et al. Longitudinal changes in Alzheimer’s-related plasma biomarkers and brain amyloid. Alzheimers Dement. Oct 2023;19(10):4335–4345. doi:10.1002/alz.13157

24. Chen S-D, Huang Y-Y, Shen X-N, et al. Longitudinal plasma phosphorylated tau 181 tracks disease progression in Alzheimer’s disease. Translational psychiatry. 2021;11(1):356. doi:10.1038/s41398-021-01476-7

25. Mattsson-Carlgren N, Janelidze S, Palmqvist S, et al. Longitudinal plasma p-tau217 is increased in early stages of Alzheimer’s disease. Brain. Dec 5 2020;143(11):3234–3241. doi:10.1093/brain/awaa286

26. Milà-Alomà M, Tosun D, Schindler SE, et al. Timing of Changes in Alzheimer’s Disease Plasma Biomarkers as Assessed by Amyloid and Tau PET Clocks. Ann Neurol. Jun 20 2025;doi:10.1002/ana.27285

27. Insel PS, Mattsson-Carlgren N, Langford O, et al. Concurrent Changes in Plasma Phosphorylated Tau 217, Tau PET, and Cognition in Preclinical Alzheimer Disease. JAMA Neurology. 2025;doi:10.1001/jamaneurol.2025.2974

28. Dyck CHv, Swanson CJ, Aisen P, et al. Lecanemab in Early Alzheimer’s Disease. New England Journal of Medicine. 2023;388(1):9-21. doi:10.1056/NEJMoa2212948

29. Sims JR, Zimmer JA, Evans CD, et al. Donanemab in Early Symptomatic Alzheimer Disease: The TRAILBLAZER-ALZ 2 Randomized Clinical Trial. Jama. Aug 8 2023;330(6):512–527. doi:10.1001/jama.2023.13239

30. Simrén J, Leuzy A, Karikari TK, et al. The diagnostic and prognostic capabilities of plasma biomarkers in Alzheimer’s disease. Alzheimer’s & dementia: the journal of the Alzheimer’s Association. 2021;17(7):1145–1156. doi:10.1002/alz.12283

31. Warmenhoven N, Salvadó G, Janelidze S, et al. A comprehensive head-to-head comparison of key plasma phosphorylated tau 217 biomarker tests. Brain. Feb 3 2025;148(2):416–431. doi:10.1093/brain/awae346

32. Rubin R. What to Know About the First FDA-Cleared Blood Test for Alzheimer Biomarkers. JAMA. 2025;doi:10.1001/jama.2025.9013

33. Jagust WJ, Landau SM. Temporal Dynamics of β-Amyloid Accumulation in Aging and Alzheimer Disease. Neurology. Mar 2 2021;96(9):e1347–e1357. doi:10.1212/wnl.0000000000011524

34. Salvadó G, Janelidze S, Bali D, et al. Plasma Phosphorylated Tau 217 to Identify Preclinical Alzheimer Disease. JAMA Neurology. 2025;doi:10.1001/jamaneurol.2025.3217

35. Kwon HS, Hwang M, Koh S-H, et al. Comparison of plasma p-tau217 and p-tau181 in predicting amyloid positivity and prognosis among Korean memory clinic patients. Scientific Reports. 2025/03/06 2025;15(1):7791. doi:10.1038/s41598-025-90232-8

36. Therriault J, Brum WS, Trudel L, et al. Blood phosphorylated tau for the diagnosis of Alzheimer’s disease: a systematic review and meta-analysis. The Lancet Neurology. 2025/09/01/2025;24(9):740-752. doi:10.1016/S1474-4422(25)00227-3

37. Pontecorvo MJ, Lu M, Burnham SC, et al. Association of Donanemab Treatment With Exploratory Plasma Biomarkers in Early Symptomatic Alzheimer Disease: A Secondary Analysis of the TRAILBLAZER-ALZ Randomized Clinical Trial. JAMA Neurology. 2022;79(12):1250–1259. doi:10.1001/jamaneurol.2022.3392

38. Horie K, Salvadó G, Koppisetti RK, et al. Plasma MTBR-tau243 biomarker identifies tau tangle pathology in Alzheimer’s disease. Nature Medicine. 2025/03/31 2025;doi:10.1038/s41591-025-03617-7

39. Schindler SE, Li Y, Li M, et al. Using Alzheimer’s disease blood tests to accelerate clinical trial enrollment. Alzheimer’s & Dementia. 2023;19(4):1175-1183. doi:10.1002/alz.12754

40. Groot C, Smith R, Stomrud E, et al. Phospho-tau with subthreshold tau-PET predicts increased tau accumulation rates in amyloid-positive individuals. Brain. Apr 19 2023;146(4):1580–1591. doi:10.1093/brain/awac329

41. Mattsson N, Cullen NC, Andreasson U, Zetterberg H, Blennow K. Association Between Longitudinal Plasma Neurofilament Light and Neurodegeneration in Patients With Alzheimer Disease. JAMA neurology. 2019;76(7):791–799. doi:10.1001/jamaneurol.2019.0765

42. Palmqvist S, Insel PS, Stomrud E, et al. Cerebrospinal fluid and plasma biomarker trajectories with increasing amyloid deposition in Alzheimer’s disease. EMBO Mol Med. Dec 2019;11(12):e11170. doi:10.15252/emmm.201911170

43. Petersen RC, Aisen PS, Beckett LA, et al. Alzheimer’s Disease Neuroimaging Initiative (ADNI): clinical characterization. Neurology. Jan 19 2010;74(3):201–9. doi:10.1212/WNL.0b013e3181cb3e25

44. Schindler SE, Petersen KK, Saef B, et al. Head-to-head comparison of leading blood tests for Alzheimer’s disease pathology. Alzheimer’s & Dementia. 2024/10/12 2024;n/a(n/a)doi:10.1002/alz.14315

45. Rowe CC, Jones G, Doré V, et al. Standardized Expression of 18F-NAV4694 and 11C-PiB β-Amyloid PET Results with the Centiloid Scale. J Nucl Med. Aug 2016;57(8):1233–7. doi:10.2967/jnumed.115.171595

46. Cho SH, Woo S, Kim C, et al. Disease progression modelling from preclinical Alzheimer’s disease (AD) to AD dementia. Scientific Reports. 2021/02/18 2021;11(1):4168. doi:10.1038/s41598-021-83585-3

