## Supplementary Materials for "Trajectories of plasma biomarkers, amyloid-beta burden and cognitive decline in Alzheimer’s disease: A Longitudinal ADNI Study"

### SUPPLEMENTARY DATA CONTENT

**Table S1.** Annualized percentage change in plasma biomarkers

**Table S2.** Comparisons of linear and quadratic model fit for the association between plasma and Aβ-PET at baseline

**Table S3.** Comparisons of linear and quadratic model fit for the association between plasma biomarkers and Aβ-PET rate of change

**Table S4.** Association between plasma rate of change of the Fujirebio Lumipulse p-tau217/A$\beta_{42}$ ratio with Aβ-PET baseline values and rate of change.

**Table S5.** Comparisons of linear and quadratic model fit for the association between baseline plasma biomarkers and Aβ-PET rate of change

**Table S6**. Progression to Aβ-PET positivity in baseline Aβ-PET negative participants in plasma A$\beta_{42}$, p-tau181, GFAP and NfL biomarkers.

**Table S7.** Association between baseline plasma levels and rate of change with ADAS13 baseline scores and rate of change.

**Table S8.** Count of missing values and outliers for plasma p-tau217, A$\beta_{42/40}$, p-tau181, GFAP and NfL in ADNI participants.

**Figure S1.** Spearman correlation matrices of plasma biomarkers, Aβ-PET, and cognitive scores across the entire cohort.

**Figure S2**. Longitudinal plasma biomarker trajectories across plasma p-tau181, GFAP and NfL.

**Figure S3**. Association between longitudinal plasma A$\beta_{42}$ and A$\beta_{40}$ biomarkers changes with Aβ-PET Centiloids accumulation over time.

**Figure S4**. Association between longitudinal plasma biomarkers changes across plasma p-tau181, NfL and GFAP with Aβ-PET Centiloids accumulation over time.

**Figure S5**. Association between baseline plasma biomarkers across plasma p-tau217, A$\beta_{42/40}$, p-tau181, NfL and GFAP and Aβ-PET Centiloids accumulation over time.

**Figure S6.** Boxplots of plasma biomarkers slopes by Aβ-PET progression status.

**Figure S7**. Association between longitudinal plasma biomarkers and MMSE rate of change.

**Figure S8.** Association between longitudinal and baseline Centiliod, MMSE and ADAS13 cognitive scores.

**Table S1.** Annualized percentage change in plasma biomarkers

| Biomarker | Assay | Full Sample | Aβ-PET positive | Aβ-PET Negative |
|---|---|---|---|---|
| ***p-tau217*** | C2N | 12.31 | 13.78 | 11.48 |
| | %C2N | 8.89 | 8.93 | 8.86 |
| | Fujirebio | 10.65 | 13.99 | 8.77 |
| | Janssen | 7.05 | 8.42 | 6.27 |
| | ALZpath | 7.78 | 8.84 | 7.18 |
| ***$A\beta_{42/40}$*** | C2N | -0.10 | 0.23 | -0.30 |
| | Fujirebio | 0.85 | 0.39 | 1.11 |
| | Roche | -0.09 | 0.06 | -0.17 |
| | Quanterix | -0.82 | -0.56 | -0.96 |
| ***p-tau181*** | Roche | 4.92 | 5.79 | 4.42 |
| | Quanterix | 4.53 | 4.91 | 4.31 |
| ***GFAP*** | Roche | 6.45 | 6.12 | 6.64 |
| | Quanterix | 6.36 | 7.70 | 5.62 |
| ***NfL*** | Roche | 5.81 | 7.43 | 4.87 |
| | Quanterix | 7.08 | 9.36 | 5.80 |

Annualized percentage change per year for plasma biomarkers. For each participant, the annualized % change was calculated by taking the relative change from baseline and last available measurements, normalized by the baseline value and divided by the time interval.

Model estimates and fit statistics from both linear and quadratic models assessing the associations between plasma biomarkers and Aβ-PET at baseline. *Notes*: The best-fitting model for each association (linear or quadratic), defined by a difference of ΔAIC and ΔBIC > 10, is indicated in bold. AIC = Akaike Information Criterion; BIC = Bayesian Information Criterion.

| | Assay | $R^2$ Linear | $R^2$ Quadratic | AIC Linear | AIC Quadratic | BIC Linear | BIC Quadratic |
|---|---|---|---|---|---|---|---|
| ***p-tau217*** | C2N | 0.51 | **0.57** | 808.57 | **762.05** | 832.18 | **789.60** |
| | % C2N | 0.60 | **0.62** | 729.52 | **709.35** | 753.11 | **736.87** |
| | Fujirebio | 0.40 | **0.48** | 882.81 | **829.21** | 906.40 | **856.73** |
| | Janssen | 0.46 | **0.48** | 848.87 | **833.37** | 872.48 | **860.91** |
| | ALZpath | 0.49 | **0.53** | 829.99 | **796.28** | 853.61 | **823.84** |
| ***$A\beta_{42/40}$*** | C2N | 0.16 | 0.18 | 1019.99 | 1012.74 | 1043.63 | 1040.32 |
| | Fujirebio | 0.19 | 0.21 | 1000.88 | 995.49 | 1024.51 | 1023.05 |
| | Roche | 0.21 | 0.21 | 981.78 | 982.07 | 1005.35 | 1009.56 |
| | Quanterix | 0.13 | 0.13 | 949.53 | 948.98 | 972.66 | 975.97 |
| ***p-tau181*** | Roche | 0.30 | **0.33** | 942.27 | **926.71** | 965.85 | **954.22** |
| | Quanterix | 0.15 | **0.19** | 935.73 | **918.07** | 958.83 | **945.01** |
| ***GFAP*** | Roche | 0.15 | 0.16 | 1011.03 | 1008.22 | 1034.61 | 1035.73 |
| | Quanterix | 0.15 | 0.15 | 939.65 | 939.65 | 962.77 | 966.61 |
| ***NfL*** | Roche | 0.04 | 0.05 | 1057.14 | 1051.08 | 1080.70 | 1078.56 |
| | Quanterix | 0.03 | 0.04 | 982.28 | 980.28 | 1005.39 | 1007.24 |

**Table S3.** Comparisons of linear and quadratic model fit for the association between plasma biomarkers and Aβ-PET rate of change

Model estimates and fit statistics from both linear and quadratic models assessing the associations

| | Assay | $R^2$ Linear | $R^2$ Quadratic | AIC Linear | AIC Quadratic | BIC Linear | BIC Quadratic |
|---|---|---|---|---|---|---|---|
| ***p-tau217*** | C2N | 0.31 | **0.49** | 948.13 | **832.99** | 971.80 | **860.61** |
| | % C2N | 0.38 | **0.49** | 910.69 | **833.81** | 934.36 | **861.43** |
| | Fujirebio | 0.26 | **0.41** | 975.96 | **887.47** | 999.63 | **915.09** |
| | Janssen | 0.29 | **0.41** | 957.54 | **887.70** | 981.22 | **915.32** |
| | ALZpath | 0.31 | **0.41** | 949.33 | **892.03** | 973.01 | **919.65** |
| ***Aβ*** $_{42/40}$ | C2N | 0.12 | 0.12 | 1043.66 | 1044.67 | 1067.33 | 1072.29 |
| | Fujirebio | 0.22 | 0.21 | 998.71 | 1000.32 | 1022.38 | 1027.93 |
| | Roche | 0.00 | 0.00 | 1088.58 | 1089.30 | 1112.24 | 1116.90 |
| | Quanterix | 0.01 | 0.01 | 1001.37 | 1003.31 | 1024.58 | 1030.40 |
| ***p-tau181*** | Roche | 0.19 | 0.22 | 898.60 | 886.94 | 921.60 | 913.76 |
| | Quanterix | 0.12 | 0.13 | 957.78 | 954.73 | 980.98 | 981.79 |
| ***GFAP*** | Roche | 0.17 | 0.18 | 1019.05 | 1013.32 | 1042.70 | 1040.92 |
| | Quanterix | 0.15 | 0.16 | 946.32 | 941.78 | 969.52 | 968.85 |
| ***NfL*** | Roche | 0.03 | 0.03 | 1080.68 | 1080.72 | 1104.35 | 1108.34 |
| | Quanterix | 0.05 | 0.05 | 988.27 | 990.06 | 1011.49 | 1017.15 |

between plasma biomarkers and Aβ-PET rates of change. *Notes*: The best-fitting model for each association (linear or quadratic), defined by a difference of ΔAIC and ΔBIC > 10, is indicated in bold. AIC = Akaike Information Criterion; BIC = Bayesian Information Criterion.

**Table S4.** Association between plasma rate of change of the Fujirebio Lumipulse p-tau217/A$\beta_{42}$ ratio with Aβ-PET baseline values and rate of change.

| | **Full sample** | | **Aβ-PET positive** | | **Aβ-PET negative** | |
|---|---|---|---|---|---|---|
| | **Estimate** | **$R^2$** | **Estimate** | **$R^2$** | **Estimate** | **$R^2$** |
| ***p-tau217/A$\beta_{42}$ rate of change*** | $0.03^{***}$ | | $0.05^{**}$ | | $0.03^{*}$ | |
| ***Aβ-PET rate of change ~ p-tau217/A$\beta_{42}$ rate of change*** | $0.43^{***}$ | 0.19 | 0.04 | 0.02 | $0.36^{***}$ | 0.12 |
| ***Aβ-PET rate of change ~ p-tau217/A$\beta_{42}$ at baseline*** | $0.37^{***}$ | 0.14 | 0.05 | 0.02 | $0.17^{**}$ | 0.02 |
| ***Aβ-PET at baseline~ p-tau217/A$\beta_{42}$ at baseline*** | $0.53^{***}$ | 0.31 | $0.39^{***}$ | 0.14 | 0.06 | 0.04 |

Model estimates and fit statistics from linear regression models assessing the associations between plasma p-tau217/A$\beta_{42}$ biomarker and cognition at baseline Aβ-PET at baseline, as well as their rates of change both in the whole group and stratified by Aβ-PET status at baseline. *Notes*: All models were adjusted for age at first plasma timepoint, years of education and sex. Uncorrected * $p < 0.05$,

**Table S5.** Comparisons of linear and quadratic model fit for the association between baseline plasma biomarkers and Aβ-PET rate of change

| | Assay | $R^2$ Linear | $R^2$ Quadratic | AIC Linear | AIC Quadratic | BIC Linear | BIC Quadratic |
|---|---|---|---|---|---|---|---|
| ***p-tau217*** | C2N | 0.26 | **0.34** | 963.45 | **920.94** | 987.06 | **948.49** |
| | % C2N | 0.29 | **0.37** | 945.43 | **904.82** | 969.03 | **932.34** |
| | Fujirebio | 0.19 | **0.30** | 998.21 | **943.81** | 1021.80 | **971.34** |
| | Janssen | 0.20 | **0.25** | 996.02 | **973.39** | 1019.62 | **1000.93** |
| | ALZpath | 0.23 | **0.30** | 984.54 | **947.55** | 1008.17 | **975.11** |
| ***Aβ$_{42/40}$*** | C2N | 0.12 | 0.12 | 1037.08 | 1037.89 | 1060.72 | 1065.47 |
| | Fujirebio | 0.17 | 0.18 | 1012.16 | 1008.19 | 1035.79 | 1035.75 |
| | Roche | 0.20 | 0.22 | 986.94 | 980.69 | 1010.50 | 1008.18 |
| | Quanterix | 0.11 | 0.12 | 955.89 | 953.66 | 979.02 | 980.64 |
| ***p-tau181*** | Roche | 0.14 | 0.16 | 1016.85 | 1007.24 | 1040.43 | 1034.75 |
| | Quanterix | 0.05 | **0.09** | 974.75 | **960.59** | 997.84 | **987.54** |
| ***GFAP*** | Roche | 0.10 | 0.10 | 1035.00 | 1034.43 | 1058.58 | 1061.93 |
| | Quanterix | 0.08 | 0.09 | 964.32 | 962.78 | 987.43 | 989.74 |
| ***NfL*** | Roche | 0.01 | 0.02 | 1066.67 | 1065.44 | 1090.23 | 1092.93 |
| | Quanterix | 0.02 | 0.02 | 988.72 | 987.31 | 1011.84 | 1014.28 |

Model estimates and fit statistics from both linear and quadratic models assessing the associations between plasma biomarkers at baseline and Aβ-PET rate of change. *Notes*: The best-fitting model for each association (linear or quadratic), defined by a difference of ΔAIC and ΔBIC > 10, is indicated in bold. AIC = Akaike Information Criterion; BIC = Bayesian Information Criterion.

**Table S6**. **Progression to Aβ-PET positivity in baseline Aβ-PET negative participants in plasma p-tau217, $A\beta_{42/40}$, $A\beta_{42}$, p-tau181, GFAP and NfL biomarkers.**

| | | Baseline | | Slope | |
|---|---|---|---|---|---|
| **Biomarker** | **Assay** | **HR** | **95% CI** | **HR** | **95% CI** |
| ***p-tau217*** | %C2N | 1.94*** | 1.62 – 2.32 | 2.45*** | 2.02 – 2.97 |
| | Fujirebio | 1.70*** | 1.41 – 2.02 | 1.90*** | 1.63 – 2.22 |
| | Janssen | 1.59*** | 1.27 – 1.99 | 1.98*** | 1.67 – 2.34 |
| | ALZpath | 1.73*** | 1.40 – 2.15 | 2.23*** | 1.81 – 2.73 |
| | C2N | 0.62** | 0.45 – 0.85 | 1.43* | 1.06 – 1.93 |
| ***$A\beta_{42/40}$*** | Fujirebio | 0.61*** | 0.51 – 0.73 | 1.81*** | 1.48 – 2.22 |
| | Roche | 0.61*** | 0.50 – 0.75 | 0.94 | 0.73 – 1.20 |
| | Quanterix | 0.70* | 0.54 – 0.91 | 0.86 | 0.66 – 1.11 |
| | C2N | 0.74 | 0.54 – 1.00 | 0.60** | 0.44 – 0.81 |
| ***$A\beta_{42}$*** | Fujirebio | 0.67* | 0.51 – 0.88 | 0.55*** | 0.41 – 0.81 |
| | Roche | 0.7* | 0.55 – 0.92 | 0.62** | 0.46 – 0.84 |
| | Quanterix | 0.78 | 0.55 – 0.92 | 0.64** | 0.47 – 0.86 |
| ***p-tau181*** | Roche | 1.25 | 0.97 – 1.62 | 1.75*** | 1.41 – 2.18 |
| | Quanterix | 0.95 | 0.68 – 1.34 | 1.66*** | 1.30 – 2.11 |
| ***GFAP*** | Roche | 1.14 | 0.85 – 1.53 | 1.35* | 1.09 – 1.67 |
| | Quanterix | 1.29 | 0.97 – 1.71 | 1.47* | 1.12 – 1.92 |
| ***NfL*** | Roche | 1.66 | 0.84 – 1.62 | 1.29 | 0.98 – 1.70 |
| | Quanterix | 1.19 | 0.87 – 1.63 | 1.34 | 0.97 – 1.85 |

Model estimates showing the hazard ratios (HRs) and 95% confidence intervals (CIs) derived from survival analyses using baseline plasma biomarkers and slopes. All models were adjusted for age at first plasma timepoint, years of education and sex. *Notes*: *$p < 0.05$, **$p < 0.005$, ***$p < 0.001$ after FDR correction for multiple comparisons across assays for each biomarker.

**Table S7.** Association between baseline plasma levels and rate of change with ADAS13 baseline scores and rate of change.

| | | | Full sample | | Aβ-PET positive | | Aβ-PET negative | |
|---|---|---|---|---|---|---|---|---|
| | **Biomarker** | **Assay** | **Estimate** | **$R^2$** | **Estimate** | **$R^2$** | **Estimate** | **$R^2$** |
| ***ADAS13 rate of change ~ plasma rate of change*** | ***p-tau217*** | C2N | $0.62^{***}$ | 0.40 | $0.58^{***}$ | 0.29 | $0.19^{*}$ | 0.09 |
| | | %C2N | $0.64^{***}$ | 0.43 | $0.63^{***}$ | 0.36 | $0.17^{*}$ | 0.09 |
| | | Fujirebio | $0.56^{***}$ | 0.34 | $0.50^{***}$ | 0.21 | 0.11 | 0.07 |
| | | Janssen | $0.61^{***}$ | 0.39 | $0.57^{***}$ | 0.29 | $0.19^{*}$ | 0.10 |
| | | ALZpath | $0.59^{***}$ | 0.36 | $0.52^{***}$ | 0.23 | $0.17^{*}$ | 0.09 |
| | ***$A\beta_{42/40}$*** | C2N | $0.20^{***}$ | 0.07 | 0.03 | -0.03 | 0.05 | 0.06 |
| | | Fujirebio | $0.31^{***}$ | 0.12 | 0.25 | 0.03 | 0.06 | 0.07 |
| | | Roche | 0.01 | 0.03 | 0.08 | -0.02 | -0.04 | 0.06 |
| | | Quanterix | $0.12^{*}$ | 0.04 | 0.18 | 0.004 | -0.07 | 0.08 |
| | ***p-tau181*** | Roche | $0.39^{***}$ | 016 | $0.26^{*}$ | 0.03 | 0.12 | 0.09 |
| | | Quanterix | $0.27^{***}$ | 0.09 | $0.22^{*}$ | 0.02 | 0.05 | 0.08 |
| | ***GFAP*** | Roche | $0.35^{***}$ | 0.13 | $0.28^{**}$ | 0.03 | 0.08 | 0.07 |
| | | Quanterix | $0.43^{***}$ | 0.17 | $0.36^{***}$ | 0.08 | 0.11 | 0.09 |
| | ***NfL*** | Roche | $0.38^{***}$ | 0.14 | $0.34^{***}$ | 0.06 | $0.32^{***}$ | 0.14 |
| | | Quanterix | $0.42^{***}$ | 0.15 | $0.38^{***}$ | 0.08 | $0.32^{***}$ | 0.15 |
| ***ADAS13 at baseline ~ plasma at baseline*** | ***p-tau217*** | C2N | $0.34^{***}$ | 0.18 | $0.30^{***}$ | 0.09 | 0.02 | 0.09 |
| | | % C2N | $0.33^{***}$ | 0.17 | $0.28^{**}$ | 0.08 | 0.002 | 0.09 |
| | | Fujirebio | $0.39^{***}$ | 0.22 | $0.43^{***}$ | 0.19 | -0.05 | 0.10 |
| | | Janssen | $0.35^{***}$ | 0.18 | $0.37^{***}$ | 0.14 | -0.06 | 0.09 |
| | | ALZpath | $0.39^{***}$ | 0.20 | $0.40^{***}$ | 0.16 | -0.08 | 0.10 |
| | ***$A\beta_{42/40}$*** | C2N | -0.14 | 0.07 | 0.04 | -0.001 | -0.09 | 0.10 |
| | | Fujirebio | -0.22 | 0.11 | -0.19 | 0.03 | -0.10 | 0.10 |
| | | Roche | -0.16 | 0.08 | 0.008 | -0.003 | -0.08 | 0.10 |
| | | Quanterix | -0.24 | 0.11 | -0.18 | 0.02 | -0.13 | 0.09 |
| | ***p-tau181*** | Roche | $0.27^{***}$ | 0.12 | $0.28^{***}$ | 0.07 | -0.04 | 0.07 |
| | | Quanterix | $0.27^{***}$ | 0.12 | $0.38^{***}$ | 0.14 | -0.08 | 0.10 |
| | ***GFAP*** | Roche | $0.18^{**}$ | 0.08 | 0.09 | 0.004 | -0.02 | 0.11 |
| | | Quanterix | $0.18^{**}$ | 0.07 | 0.15 | 0.01 | -0.14 | 0.10 |
| | ***NfL*** | Roche | $0.21^{***}$ | 0.09 | $0.24^{*}$ | 0.04 | 0.11 | 0.10 |
| | | Quanterix | $0.20^{***}$ | 0.08 | $0.25^{*}$ | 0.05 | 0.05 | 0.07 |

Model estimates and fit statistics from linear regression models assessing the associations between plasma biomarkers and ADAS13 at baseline, as well as their rates of change. *Notes*: $^{*}p < 0.05$, $^{**}p < 0.005$, $^{***}p < 0.001$ after FDR correction for multiple comparisons across assays for each biomarker. ADAS13 = Alzheimer's Disease Assessment Scale-Cognitive Subscale 13

**Table S8.** Count of missing values and outliers for plasma p-tau217, $A\beta_{42/40}$, p-tau181, GFAP and NfL in ADNI participants.

| Assay | Number of participants with missing values | Number of outlier values |
|---|---|---|
| ***p-tau217*** | | |
| C2N | 26 | 2 |
| % C2N | 28 | 3 |
| Fujirebio | 26 | 6 |
| Janssen | 27 | 2 |
| ALZpath | 30 | 2 |
| ***$A\beta_{42/40}$*** | | |
| C2N | 29 | 0 |
| Fujirebio | 26 | 0 |
| Roche | 32 | 2 |
| Quanterix | 55 | 1 |
| ***p-tau181*** | | |
| Roche | 31 | 2 |
| Quanterix | 55 | 6 |
| ***GFAP*** | | |
| Roche | 31 | 4 |
| Quanterix | 54 | 0 |
| ***NfL*** | | |
| Roche | 29 | 2 |
| Quanterix | 54 | 2 |

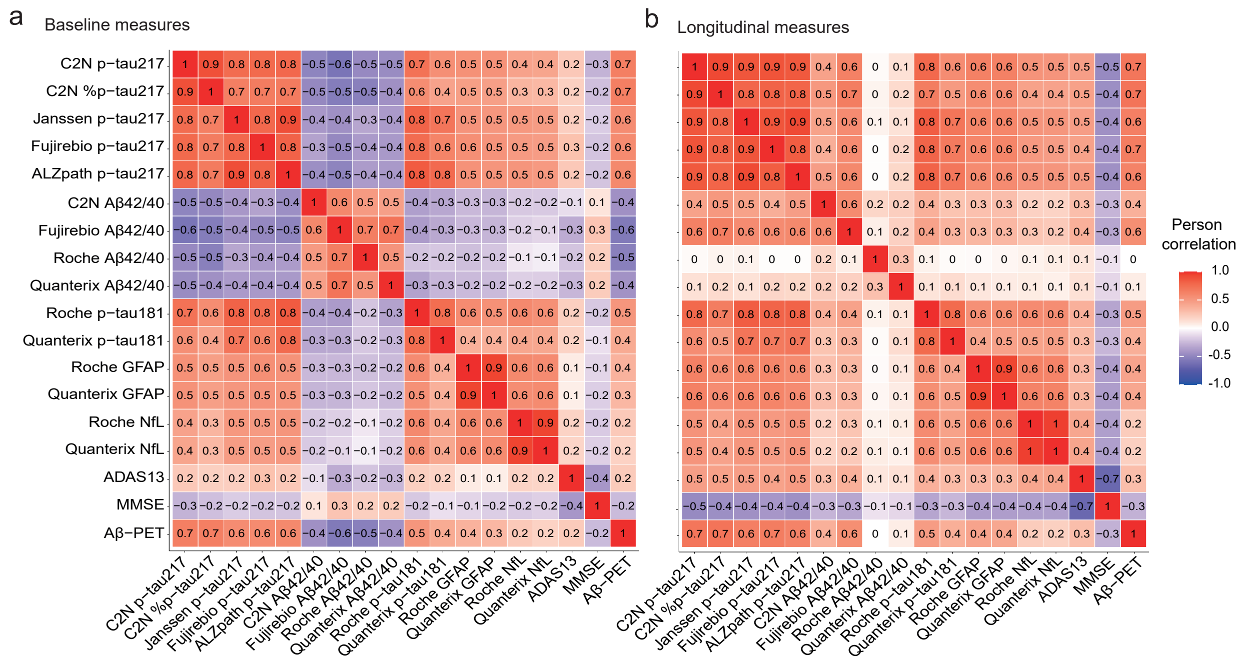


**Figure S1. Spearman correlation matrices of plasma biomarkers,** Aβ-PET**, and cognitive scores across the entire cohort. Panel A)** Correlation matrix at baseline. **Panel B)** Correlation matrix of biomarker slopes. Color intensity reflects the strength of the correlation: deeper red indicates stronger absolute correlations, while deeper blue indicates weaker absolute correlations.

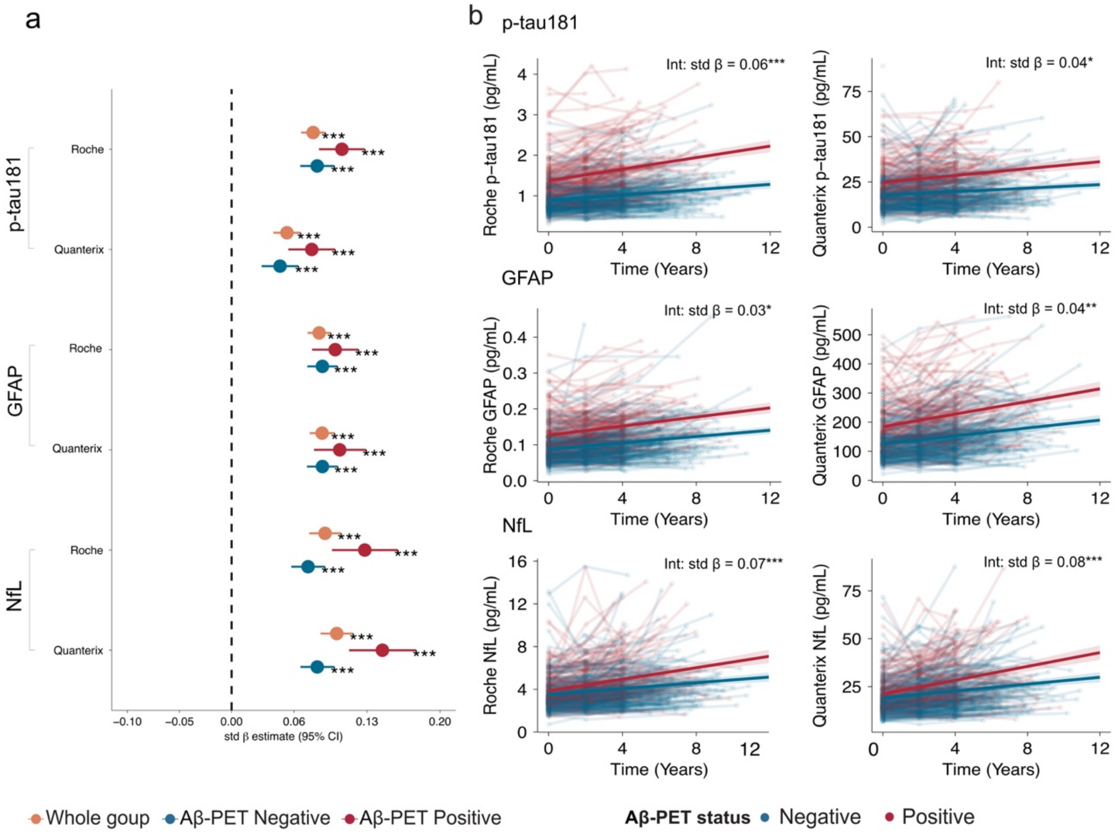


**Figure S2**. **Longitudinal plasma biomarker trajectories across plasma p-tau181, GFAP and NfL. Panel A)** Standardized regression coefficients (std β) and 95% confidence interval (CI) from linear-mixed effect models assessing plasma levels over time (plasma ~ time) **A)**. across the whole group (orange) and across the Aβ-PET negative (blue) and positive (red) groups. **B).** Different plasma biomarkers overtime with the interaction between time and Aβ status at baseline. The *x* axis shows time from first plasma biomarker sample. *Notes*: All models were adjusted for age at first plasma timepoint, years of education and sex. **p* < 0.05, ***p* <0.005, ****p* <0.001.

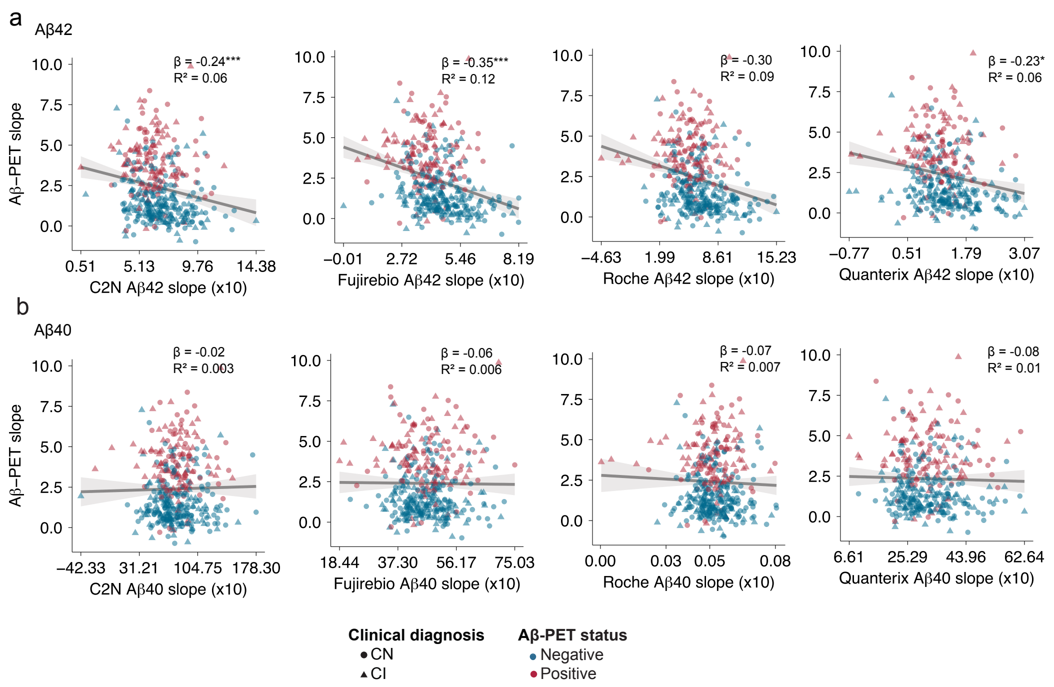


**Figure S3**. **Association between longitudinal plasma Aβ42 and Aβ40 biomarkers changes with Aβ-PET Centiloids accumulation over time.** Scatterplots illustrating the association between plasma Aβ42 (Panel A) and Aβ40 (Panel B) levels rate of change (x-axis) and the Centiloid rate of change (y-axis) in the whole group. *Notes*: Roche Aβ40 is expressed in ng/mL, while all other biomarkers are in pg/mL.*$p < 0.05$, **$p < 0.005$, ***$p < 0.001$.

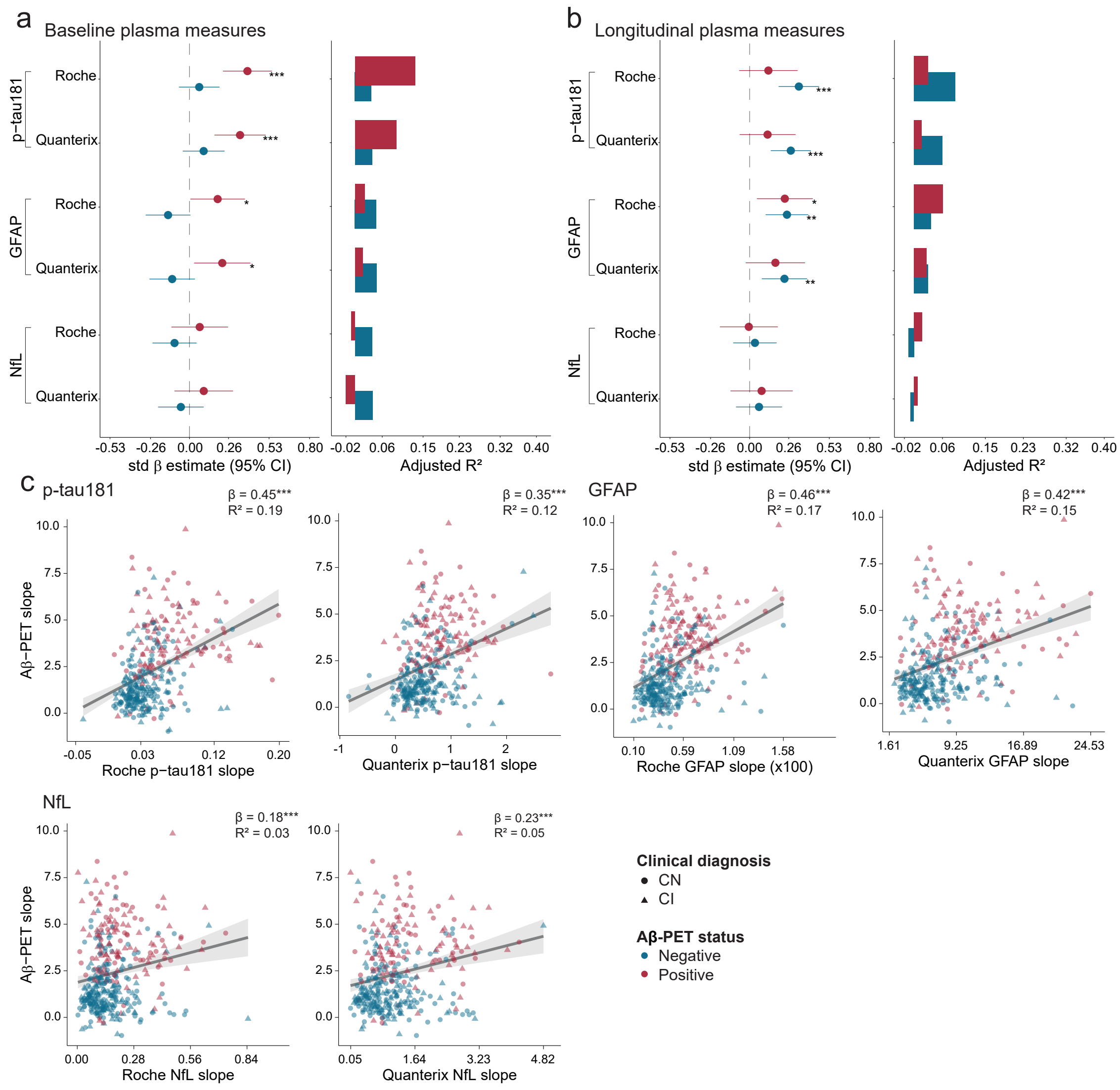


**Figure S4**. **Association between longitudinal plasma biomarkers changes across plasma p-tau181, NfL and GFAP with Aβ-PET Centiloids accumulation over time. Panel A).** Forest plots (left) of std β and 95% CIs and bar plots (right) of adjusted $R^2$ for the Aβ-PET negative group (blue) and the Aβ-PET positive group (red), derived from linear regression models of baseline plasma biomarkers association with baseline Aβ-PET. **Panel B).** Similar plots as in A derived from linear regression models of plasma biomarkers and plasma biomarkers change and Aβ-PET change. **Panel C).** Scatterplots illustrating the association between plasma levels rate of change (x-axis) and the Centiloid rate of change (y-axis) in the whole group. *Notes*: All models were adjusted for age at first plasma timepoint, years of education and sex. *$p < 0.05$, **$p < 0.005$, ***$p < 0.001$ after FDR correction for multiple comparisons across assays for each biomarker.

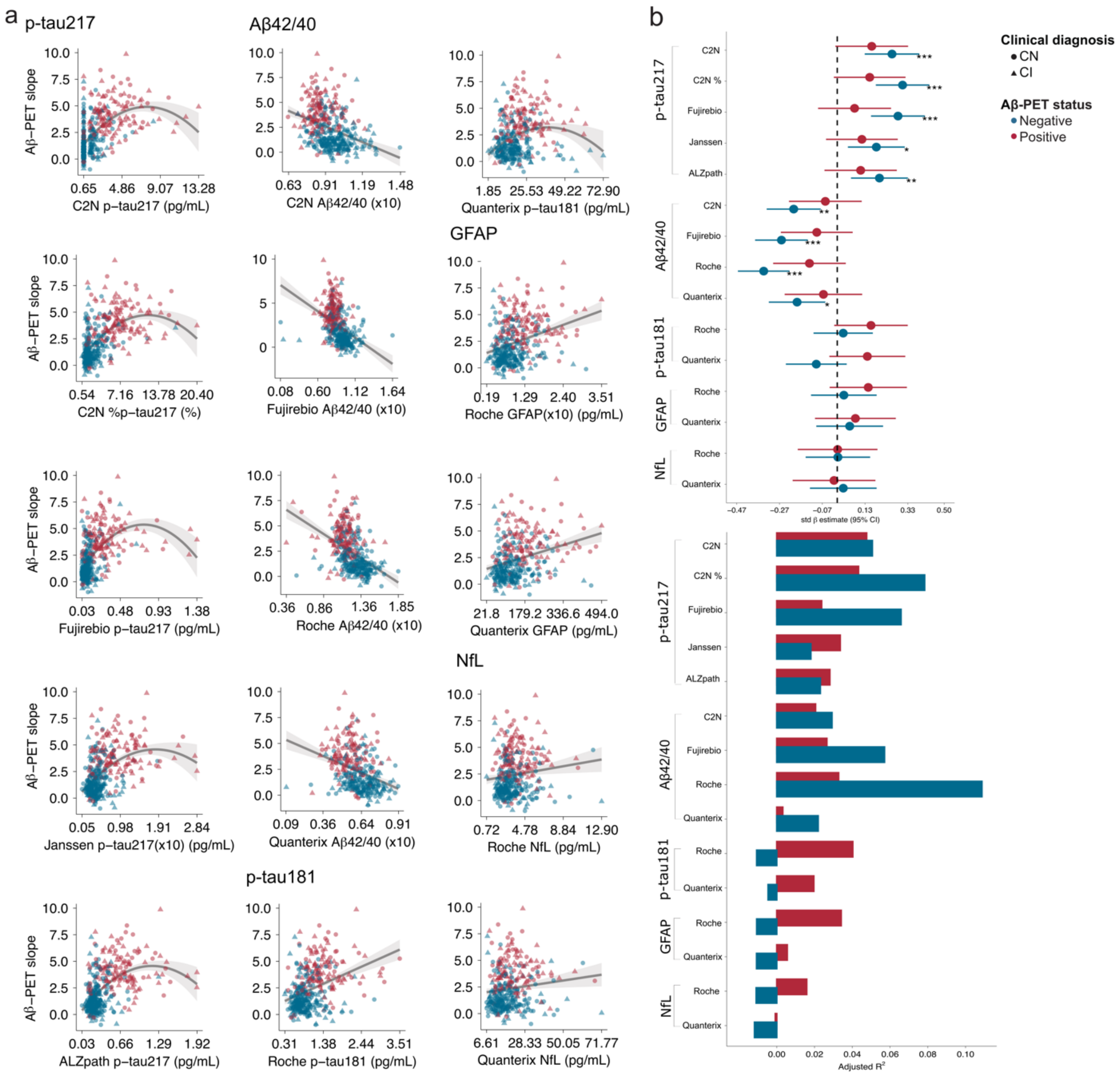


**Figure S5**. **Association between baseline plasma biomarkers across plasma p-tau217, Aβ42/40, p-tau181, NfL and GFAP and Aβ-PET Centiloids accumulation over time. Panel A).** Scatterplots illustrating the association between baseline plasma levels (x-axis) and the Centiloid rate of change (y-axis) in the whole group with quadratic and linear regression applied where appropriate. **Panel B).** Forest plots (left) of the standardized regression coefficients (std β) and 95% confidence intervals (CIs) and bar plots (right) of adjusted $R^2$ for the Aβ-PET negative group (blue) and the Aβ-PET positive group (red), derived from linear regression models of baseline plasma biomarkers and Aβ-PET change. *Notes*: All models were adjusted for age at first plasma timepoint, years of education and sex. *$p$ < 0.05, **$p$ <0.005, ***$p$ <0.001 after FDR correction for multiple comparisons across assays for each biomarker.

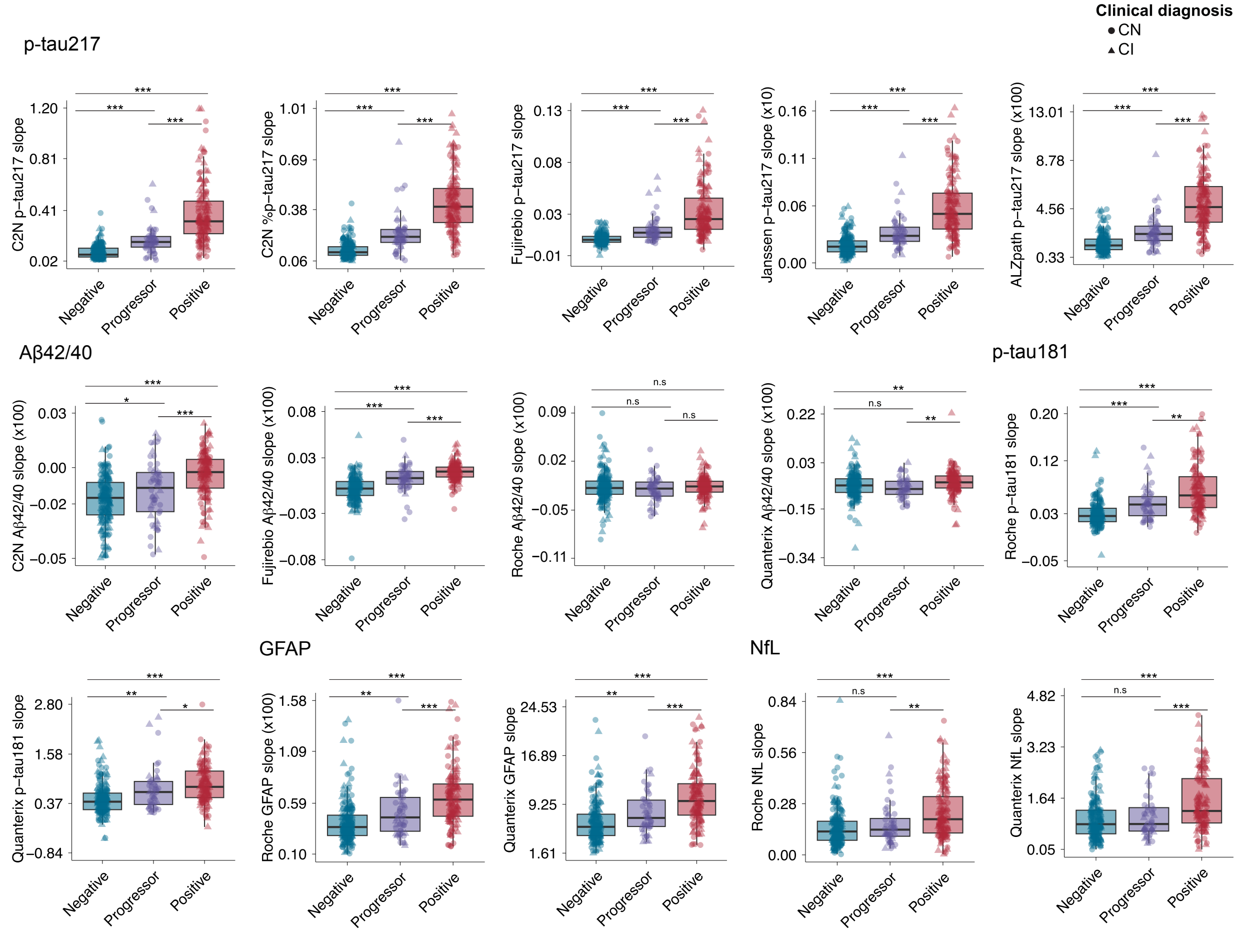


**Figure S6. Boxplots of longitudinal plasma biomarkers changes by Aβ-PET progression status.** Boxplots of plasma p-tau217, Aβ$_{42/40}$, p-tau181, GFAP and NfL rate of change across various assays were compared between Aβ-PET negative stable participants **(blue)** (individuals who remained negative across all PET scans); Aβ-PET progressors **(purple)** (individuals who were Aβ-PET negative at baseline and progressed to positive at follow-up scans); and Aβ-PET positive stable participants **(red),** individuals who remained positive across all available scans. Group comparisons were performed using the Kruskal-Wallis test followed by Dunn's post-hoc test for multiple comparisons **p* < 0.05, ***p* <0.005, ****p* <0.001

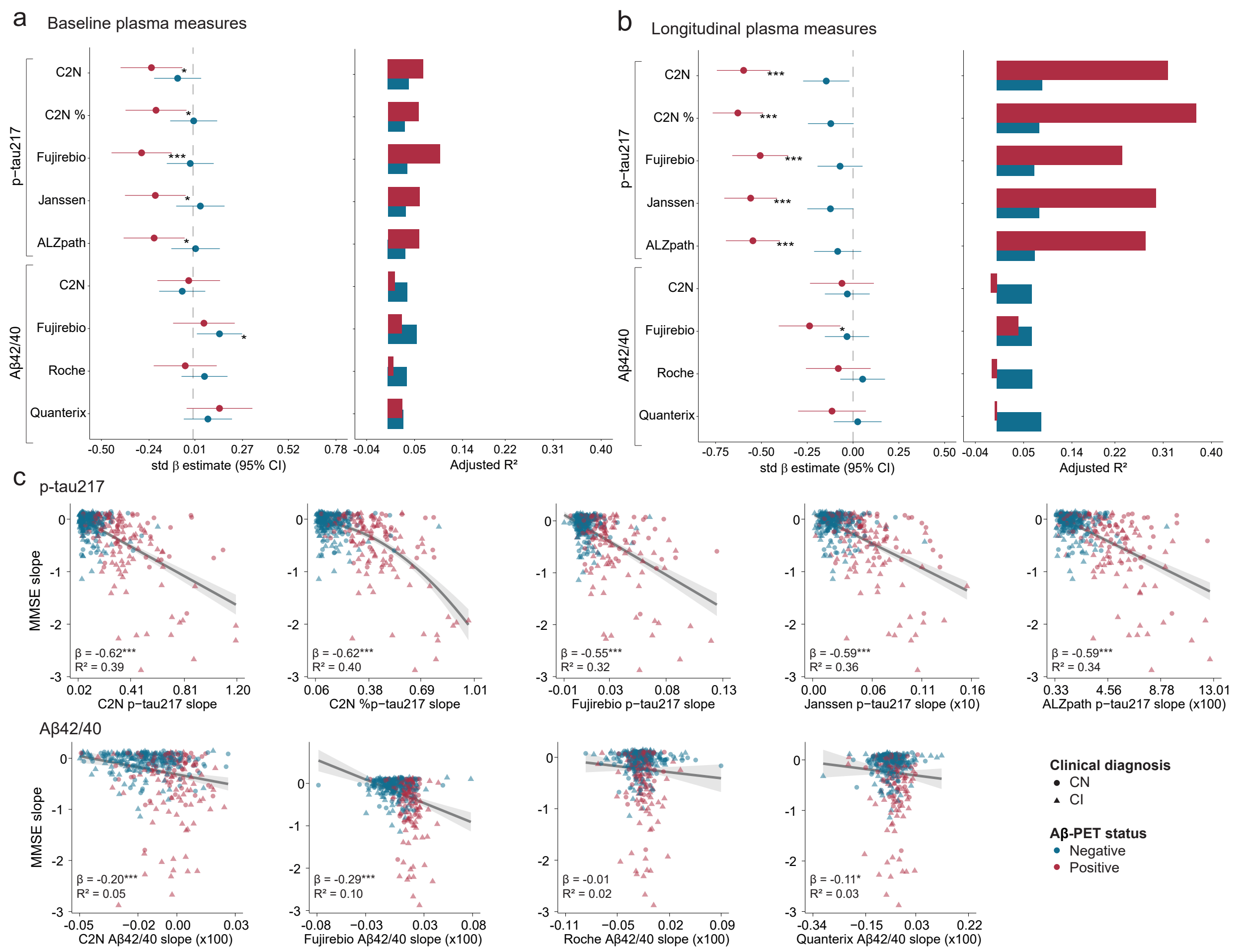


**Figure S7. Association between longitudinal plasma biomarkers and MMSE rate of change. A)** Forest plots (left) of std β and 95% CIs and bar plots (right) of adjusted $R^2$ for the Aβ-PET negative group (blue) and the Aβ-PET positive group (red), derived from linear regression models of baseline plasma biomarkers association with baseline ADAS13 score. **B)** Similar plots as in A derived from linear regression models of plasma biomarkers change and change in MMSE. **C)** Scatterplots showing the rate of change in plasma levels (x-axis) and ADAS13 (y-axis) in the whole group with quadratic (% C2N p-tau217) and linear regression applied where appropriate. The std β, adj $R^2$ and p-values from linear model for %C2N p-tau217 is reported. All models were adjusted for age at first plasma timepoint, years of education and sex. MMSE: Mini-Mental State Examination. *$p < 0.05$, **$p < 0.005$, ***$p < 0.001$ after FDR correction for multiple comparisons across assays for each biomarker.

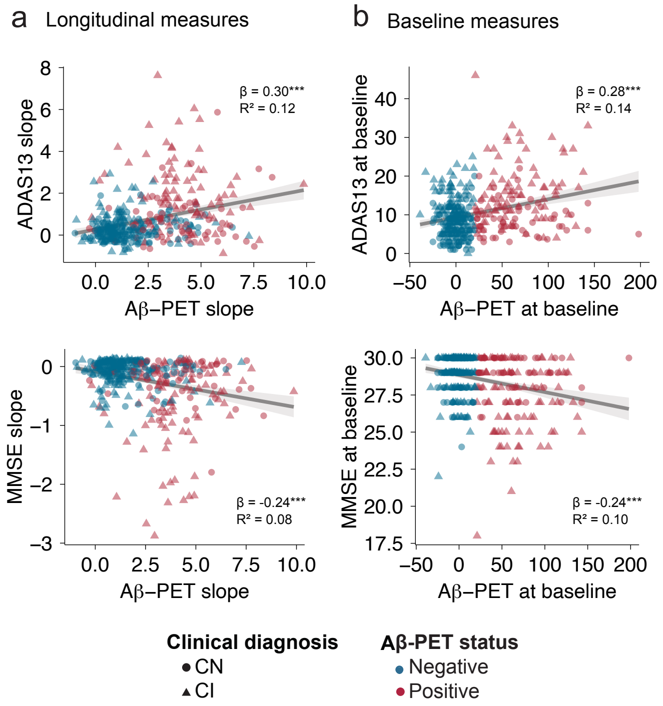


**Figure S8. Association between longitudinal and baseline Centiloid, MMSE and ADAS13 cognitive scores. Panel A).** scatterplots illustrating the linear association between Centiliod rate of change as observed on Aβ-PET (x-axis), ADAS13 and MMSE rate of change (y-axis) in the whole group. **Panel B).** scatterplots illustrating the linear association between baseline Centiliod levels as observed on Aβ-PET (x-axis), ADAS13 and MMSE baseline scores (y-axis) in the whole group. *Notes*: All models were adjusted for age at first plasma timepoint, years of education and sex.. ADAS13 = Alzheimer's Disease Assessment Scale-Cognitive Subscale 13. MMSE = Mini Mental State Examination. *$p < 0.05$, **$p < 0.005$, ***$p < 0.001$.
